# Meta Mesh Ontology: A Transformative Approach to Mental Health Knowledge Integration

**DOI:** 10.1101/2025.11.04.25339387

**Authors:** Rogério Blitz, Maike Richter, André Kerber, Ramona Leenings, Sylvia Thun, Nils Opel

## Abstract

Psychiatric diagnostics struggle to capture the complexity of mental disorders, thereby hampering individualized treatment and personalized psychiatry. Conventional classification systems like DSM-5 and ICD-11 impose rigid categories that inadequately reflect multifactorial, dynamic, and comorbid conditions. To address this limitation, alternative frameworks such as dimensional models and domain ontologies for structured knowledge representation have emerged, yet they often remain isolated and non-interoperable. We present the Meta Mesh Ontology (MMO) as an integrative framework unifying heterogeneous psychiatric knowledge bases through a modular, BFO-compliant structure. The MMO ensures semantic and structural consistency across biological, psychological, and social domains, enabling harmonized data representation and cross-domain reasoning. Its interoperability with established ontologies and informatics standards facilitates integration of multimodal data from clinical, environmental, and sensor sources. By bridging diverse taxonomies such as HiTOP and RDoC, the MMO establishes a scalable foundation for data harmonization and personalized, multidimensional diagnostics in mental health research and practice.

## 1 Introduction

Developments in digital psychiatry highlight the need for structured, interoperable, and context-sensitive data frameworks to better capture the complex interplay of biological, psychological, and social factors in mental disorders. The Meta Mesh Ontology (MMO) presented in this paper addresses this challenge by providing a foundation for integrated, semantically coherent, and ethically responsible data representation in mental health care.

### 1.1 Challenges in Psychiatric Diagnostics

Psychiatric diagnostics face the challenging task of comprehensively capturing the multifaceted and often intertwined factors of mental disorders. This involves not only the precise identification and classification of symptoms but also the consideration of biological, social, and psychological influences that shape the individual clinical picture [1-5]. Despite their complexity, current diagnostic approaches remain incomplete, lacking consistent methods to account for socioeconomic parameters that significantly affect patients’ living conditions and overall well-being. In the context of personalized psychiatry, the goal is to develop individualized treatment approaches based on precise and differentiated diagnostics that are optimally tailored to the specific needs and living conditions of each patient. This aims to maximize therapeutic efficacy and sustainably improve the prognosis for affected individuals. Traditional classification systems, such as the Diagnostic and Statistical Manual of Mental Disorders, Fifth Edition (DSM-5) [6,7] and the International Classification of Diseases, 11th Revision (ICD-11) [8-10], which focus on symptomatic criteria, often reach their limitations in this regard. Their structure is designed to capture mental disorders within rigid, predefined categories, which frequently fail to adequately reflect the diversity and dynamics of these conditions [11]. This limitation becomes particularly evident in patients with complex symptom constellations and comorbidities, where diagnostic precision suffers, making tailored therapeutic approaches more difficult. Furthermore, the high variability of symptoms among different patients presents a significant challenge. Even with identical diagnoses, mental disorders can manifest in highly diverse ways and be subjectively experienced differently, complicating standardized diagnostics - for example, Borderline Personality Disorder can encompass up to 256 possible symptom combinations. This diversity is influenced by cultural, social, and biological factors that are often not sufficiently considered in the diagnostic process. Incorporating individual life circumstances and patients’ biographical histories into both the diagnostic process and the formulation process is therefore essential for ensuring a holistic assessment of both mental and physical health aspects [12].

### 1.2 Integration of Alternative Knowledge Representation Systems

Alternative knowledge representation systems, such as the Hierarchical Taxonomy of Psychopathology (HiTOP) [13,14] and the Research Domain Criteria (RDoC) framework [15], have been developed to capture the multi-causal and interactive nature of mental disorders. These systems offer a more dynamic perspective that extends beyond traditional, rigid diagnostic typologies, considering the wide range of symptoms and their interactions [16]. However, a key limitation is that these systems are often applied in isolation and remain predominantly symptom-focused, thus overlooking crucial contextual information, including risk factors and social determinants of mental health. The lack of standardized and interoperable codifications significantly hampers the use of these alternative knowledge representation systems in both clinical practice and research [17,18]. Unified data formats and classification schemes are required to effectively integrate and consolidate data, enabling coherent data harmonization. The absence of such uniform standards results in the loss of valuable information, either by not being systematically captured or shared across different institutions and studies. A comprehensive networking and integration of heterogeneous data systems is currently only partially possible, which severely hinders the development of truly personalized and dynamic diagnostics because disconnected and non-harmonized data sources do not allow the correlation of biological, psychological, and social factors, thereby preventing a holistic understanding of individual patient trajectories. These challenges are not only technical but also organizational, as many clinics and research institutions utilize different systems and approaches. This lack of integration not only limits diagnostic precision but also the development of individualized treatment strategies and the understanding of underlying biopsychosocial patterns. This paper addresses these challenges by introducing the Meta Mesh Ontology (MMO), a new integrative framework designed to harmonize heterogeneous data sources and enable the semantic interconnection of clinical, biological, environmental, and social information. The MMO provides the methodological foundation for a unified, scalable, and ethically grounded representation of psychiatric knowledge. Building on this, the foundation is established for the application of artificial intelligence in psychiatry. Structured and semantically interoperable data constitute a central prerequisite for developing explainable, ethically responsible, and clinically applicable AI systems that can support diagnosis, prognosis, and personalized treatment planning.

## 2 Results

### 2.1 Introduction to the Meta Mesh Ontology

To address the outlined shortcomings, we introduce the Meta Mesh Ontology (MMO) [19], a unifying framework designed to integrate heterogeneous knowledge bases within a coherent and interoperable structure. The MMO functions as a meta-ontology, acting as middleware between heterogeneous knowledge systems. In this capacity, it serves as an intermediary between various ontologies and knowledge representations.

**Figure 1.**
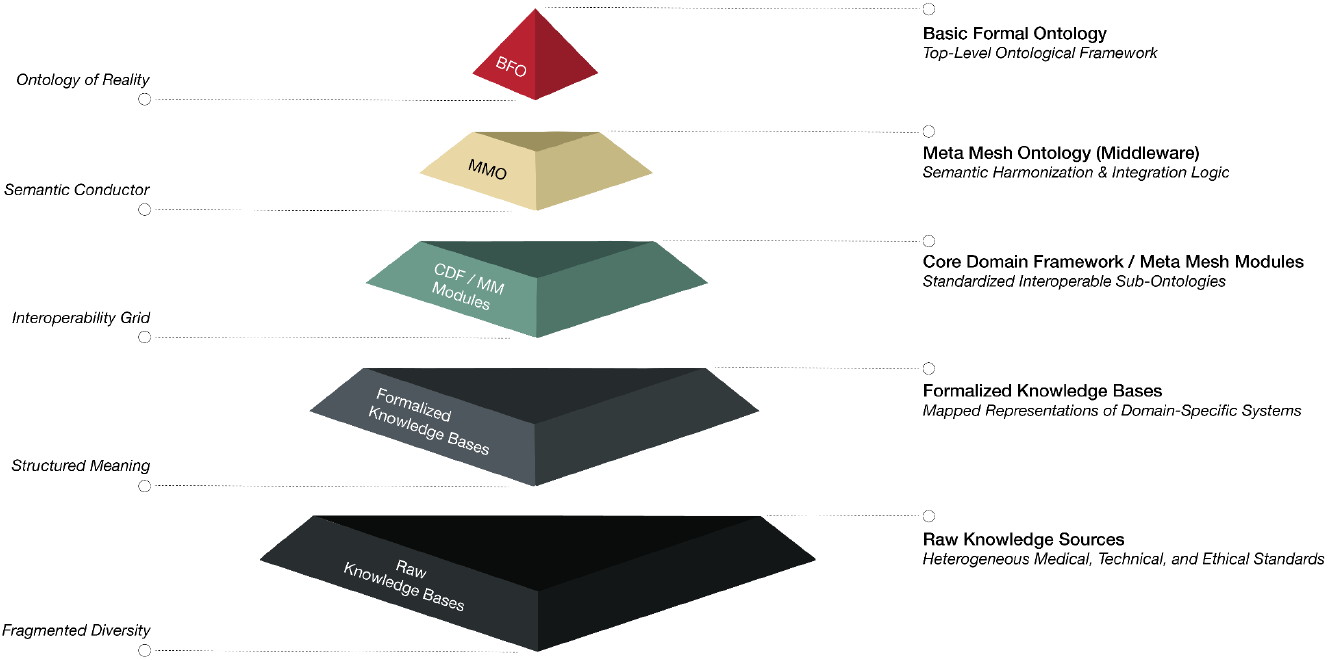
Layered architecture of the Meta Mesh Ontology (MMO), illustrating its role as a middleware framework between foundational ontology (BFO) and heterogeneous knowledge sources via modular, interoperable sub-ontologies.

Through abstraction, standardization, harmonization, and modularity, it provides a higher-level structure that allows different knowledge systems to be integrated as sub-ontologies without compromising their specific characteristics and definitions. It establishes a unified semantic interface to ensure interoperability between different knowledge representations, thereby serving as a connecting layer between heterogeneous ontologies. The MMO incorporates both syntactic and semantic rules that enable precise mapping and transformation of entities between ontologies. Additionally, it provides specific navigation mechanisms and transformation rules to ensure consistent exploration and modification of ontology elements and associated data. Its modular design allows for flexible adaptation to new knowledge domains while ensuring that integration requirements remain valid even in dynamically evolving knowledge areas. Strict integration guidelines ensure that new sub-ontologies can be seamlessly added or existing structures updated without destabilizing the overall framework. By employing mapping rules and refinement relationships, the meta-ontology supports detailed specification of concept relationships. Furthermore, it offers mechanisms for consistency monitoring and validation of newly integrated modules, ensuring a coherent knowledge base and enabling early detection of potential semantic conflicts. This promotes the quality and efficiency of data integration while reducing the effort required for maintaining and adapting the ontology system.

### 2.2 Structure and Modular Architecture

Modeling and structuring knowledge is a central challenge in knowledge representation. Based on a unified philosophical foundation, the goal is to enable a precise and coherent modeling of reality. To achieve this, the Meta Mesh Ontology (MMO) utilizes the Basic Formal Ontology (BFO) as its upper-level ontology [20,21]. The BFO is characterized by a highly axiomatized and formal structure, dividing reality into two fundamental categories: Entities that persist over time without losing their identity [22], as well as processes or events that exist within time [23]. This structure is essential for medical knowledge representation, as it allows for the depiction of both states and processes. Building upon this, the MMO establishes a robust foundation, ensuring both structural and semantic consistency. In integrating medical, psychological, and social aspects, the MMO provides a structured framework of concepts and relationships, that enables the efficient unification of existing knowledge systems. Based on established meta-ontological design standards, the MMO was developed according to the following architectural principles to ensure cross-domain interoperability, scalability, and semantic consistency.

**Table 1.** Key Architectural Principles of the Meta Mesh Ontology Design.

| Key Architectural Principles |  |
| --- | --- |
| Formal Consistency | Minimization of semantic inconsistencies through a strict axiomatic foundation. |
| Modularity and Reusability | Supports reusability and integration through a meta-level structure. |
| Cross-Domain Applicability | Provides a general structure for consistent representation across various disciplines. |
| Interoperability | Enhances interoperability between different systems and platforms through standardized concepts and relations. |
| Ontology Alignment and Mapping Support | Facilitates the integration of knowledge bases from different disciplines. |
| Automated Reasoning Capabilities | Supports automated reasoning mechanisms to derive new insights from existing data and aid decision-making. |
| Usability | Transparent ontology structure to support interdisciplinary usability, improving understanding and acceptance across different fields. |
| Transparency and Traceability | An axiomatic foundation ensures clear documentation of the origin and rationale behind conceptualization, improving transparency and traceability of knowledge representations. |

The modular architecture enables the flexible integration of sub-ontologies and their adaptation to specific application areas. Building on established ontologies and terminologies, this design ensures semantic consistency, compatibility, and seamless interoperability across knowledge systems. To support semantic interoperability across heterogeneous knowledge domains, the Meta Mesh Ontology (MMO) employs a Core Domain Framework (CDF) [24] as its top-level domain architecture. The CDF defines eight disjoint but extensible superdomains - Knowledge, Culture, Health, Nature, Technology, Economy, Governance, and Society - each representing a generalizable and semantically coherent area of human knowledge. These domains serve as anchor points for modular integration and ensure consistent alignment with the Basic Formal Ontology (BFO). The derivation process followed a multi-stage synthesis of ontological, epistemological, and classificatory perspectives and is documented in detail in the supplementary material. This structured domain scaffold ensures that all domain-specific modules within the MMO can be semantically positioned in a consistent and interoperable way. To illustrate how the Core

Domain Framework (CDF) integrates into the overall ontology structure, a schematic architectural overview is provided below.

**Figure 2.**
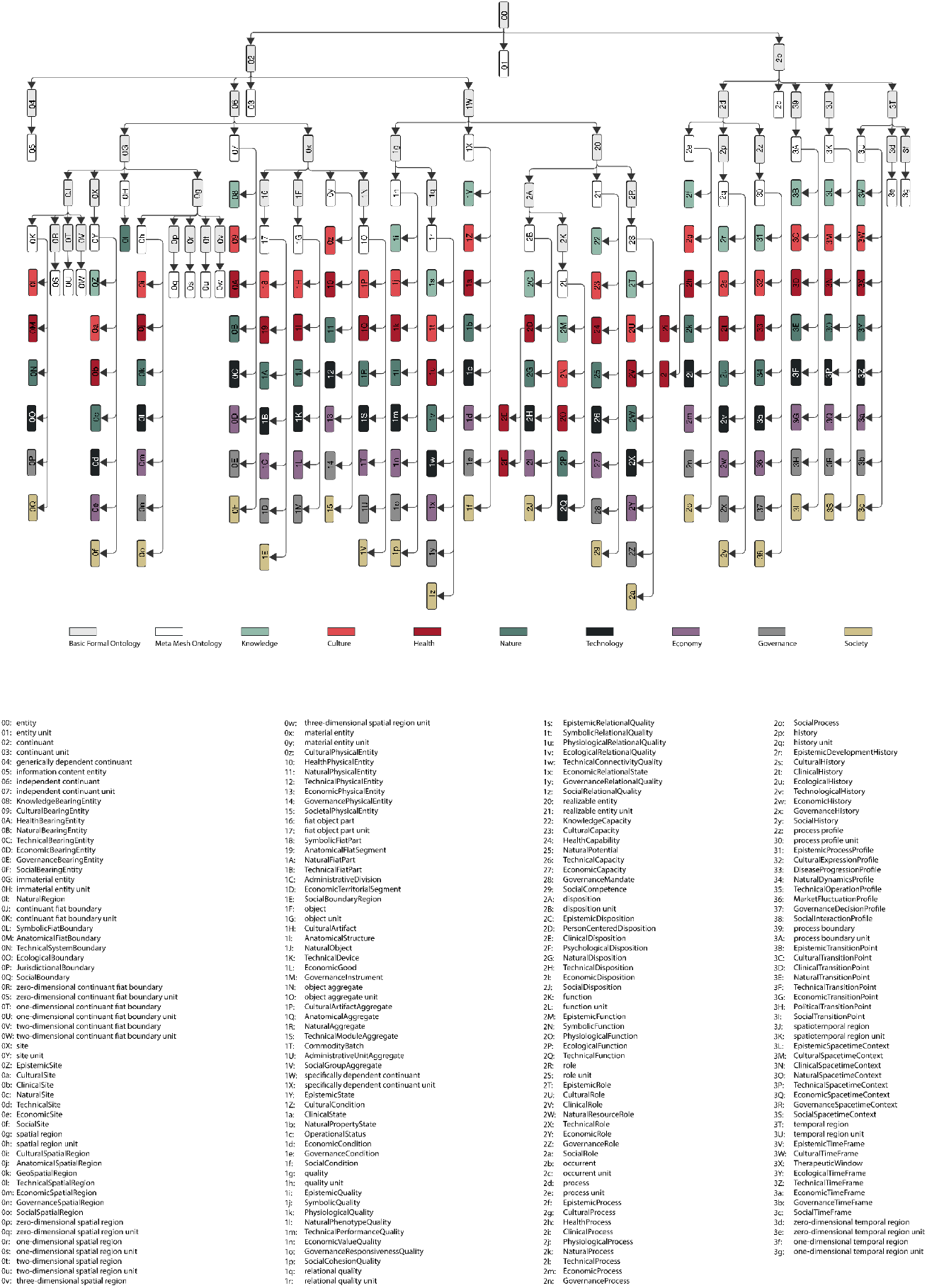
Schematic overview of the MMO architecture. The illustration shows the top-level alignment with the Basic Formal Ontology (BFO) and the integration of the Core Domain Framework (CDF) classes as intermediary structures that connect domain-specific modules to the foundational layer. The upper part illustrates the hierarchical structure and interrelations of the MMO and CDF classes. The lower part lists the corresponding class names. Each class is assigned a unique identifier encoded in base62 notation, allowing the abbreviations in the figure to be matched to their respective class names.

### 2.3 Interoperability and Technical Integration

The Meta Mesh Ontology (MMO) supports the harmonization of diverse knowledge domains by providing a set of BFO-compilant classes. For each BFO class, the MMO specifies a corresponding MMO subclass that acts as a semantic mediator and alignment layer. These MMO subclasses serve as supercategories to structure domain-specific concepts and to establish a coherent organization of knowledge domains. In this way, interoperability between technical systems, sensor technology, and other relevant areas - such as health informatics, clinical decision support systems, and biomedical data analysis - is seamlessly ensured. The decision to adopt BFO-compliant categories is intended to enhance both comprehensibility and traceability for users and developers while also facilitating the future integration of new knowledge domains and standards. This results in a framework that can flexibly respond to the rapidly evolving demands of medicine and technology without losing expressiveness or coherence. To promote interoperability and the effective utilization of heterogeneous data sources, the MMO provides a means to integrate data from areas such as telemedicine, imaging diagnostics, and sensor technology into a unified structure. This is particularly relevant for establishing continuous patient monitoring and supporting precise, data-driven decisions in clinical settings. The MMO fosters scalability and supports future developments, including the incorporation of new sensor and diagnostic techniques as well as integration into research networks. By ensuring the consistent collection and semantic analysis of multimodal data, it enables a broad range of applications, such as early disease detection, identification of individual health risks, and the optimization of prevention strategies. Ultimately, the ontology is intended to serve as a foundation for sustainable knowledge management, facilitating communication between research, clinical practice, and technology through clearly defined terms and relationships while ensuring high standards in data quality and consistency. The precise differentiation and modularization of ontology components allow for flexible adaptation to specific use cases and facilitate future developments.

### 2.4 Use Case: Enabling Integrated Mental Health Care through the Meta Mesh Ontology

#### 2.4.1 Background

Mental health care frequently faces the challenge of fragmented data landscapes, where critical information is distributed across electronic health records (EHRs), social determinants of health, sensor data (from wearables and smart devices), environmental parameters, and behavioral assessments. This fragmentation hinders accurate diagnostics, risk stratification, and the application of AI-based methods within clinical workflows, limiting clinicians’ ability to deliver integrated and personalized care. To illustrate the practical application of the MMO, a patient case is presented. This case demonstrates the semantic integration of heterogeneous clinical and contextual data in mental health care.

#### 2.4.2 Case Description

A 38-year-old female patient presents at a psychiatric outpatient clinic reporting sleep disturbances, anhedonia, and fatigue. Her electronic health record contains isolated documentation of previous depressive episodes and somatic complaints. Relevant social data - such as unemployment, single parenting, and housing insecurity - may exist in social service records, while wearable device data would reveal reduced physical activity and fragmented sleep patterns. In conventional clinical workflows, these data remain siloed, inconsistently coded, and lack semantic interoperability. This fragmentation hinders their integration into clinical decision-making processes and constrains the effective use of AI-based analytic tools.

#### 2.4.3 Application of the Meta Mesh Ontology

The Meta Mesh Ontology (MMO) was applied in this case to demonstrate the integration of heterogeneous data sources within digital mental health care. For this purpose, the MMO was implemented within a psychiatric outpatient environment and connected to existing electronic health records (EHR), primary systems, and data management platforms through standardized interoperability protocols such as HL7 FHIR and RDF/XML. The ontology layer functioned as a semantic middleware, integrating clinical documentation, social data, and wearable sensor information into a unified, machine-readable structure. In total, 50 heterogeneous data points from medical, social, and behavioral domains were incorporated, including:

- Semantically normalized across domains, including medical symptoms, social determinants, trauma history, sensor-based data, biomarkers, molecular data, environmental parameters, behavioral patterns, and care trajectories.
- Mapped to international medical standards, including Systematized Nomenclature of Medicine - Clinical Terms (SNOMED CT) [25,26], International Classification of Diseases, Version 10 (ICD-10) [27], Logical Observation Identifier Names and Codes (LOINC) [28,29], Unified Medical Language System (UMLS) [30,31], Social Determinants of Health Ontology (SOHO) [32], Adverse Childhood Experiences Ontology (ACESO) [33], Gene Ontology (GO) [34], Hierarchical Taxonomy of Psychopathology (HiTOP) [10], and Sustainable cities and communities - Indicators for city services and quality of life (ISO 37120) [35], utilizing the modular architecture of the MMO and the Core Domain Framework (CDF).
- Transformed into a machine-readable, FAIR-compliant patient representation, enabling seamless use within clinical dashboards and AI-based analytical pipelines.

By applying the MMO, these previously distributed data points were linked within a unified semantic framework, allowing for consistent interpretation across clinical and technical systems. The following use case visualization exemplifies this integration by depicting the full spectrum of data points aligned under the Core Domain Framework (CDF). Detailed information on each of the 50 data points - including domain assignment and semantic encoding - is provided in tabular form in the supplementary materials.

**Figure 3.**
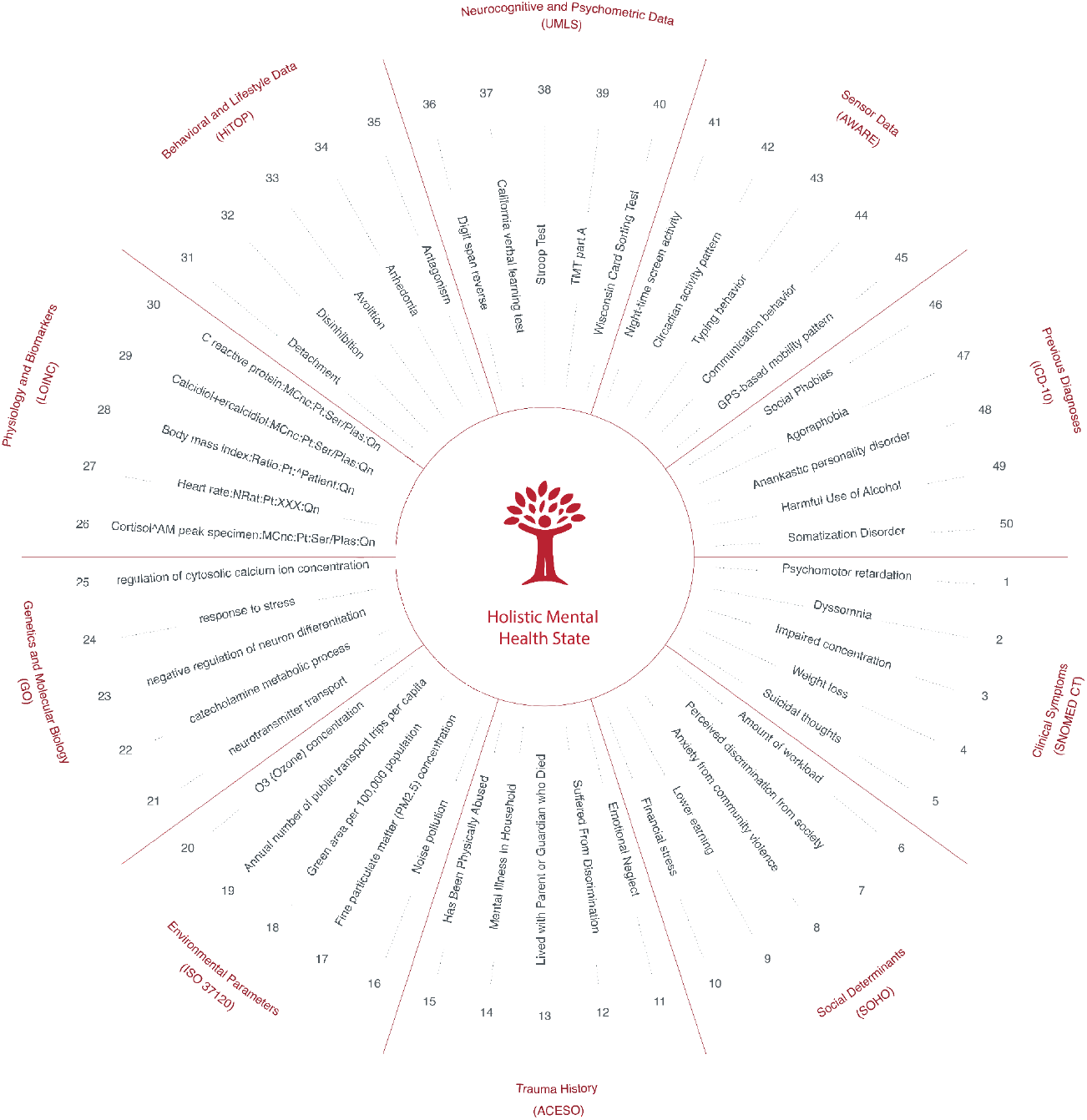
Integrated representation of 50 heterogeneous data points using the Meta Mesh Ontology (MMO). The illustration demonstrates how clinical, psychological, environmental, and social information can be unified through domain-level anchoring within the Core Domain Framework (CDF).

#### 2.4.4 Impact on Clinical Care

The application of the MMO in this use case demonstrates several critical impacts:

- Holistic Patient View: Clinicians can access an integrated, interoperable patient profile that consolidates medical, social, and behavioral data, supporting comprehensive clinical assessment. In the presented case, this profile may then lead to an intervention that primarily involves psychosocial or social work-based interventions. The integration of social determinants and behavioral data in this case may reveal that the patient’s depressive symptoms are closely linked to chronic occupational stress and social isolation. This insight can prompt a coordinated response involving psychotherapy, workplace mediation, and community-based support services.
- Enhanced AI Utility: AI models can utilize the semantically unified data structure to predict the risk of chronic depression and recommend individualized care plans with improved explainability and fairness.
- Inclusion of Social Determinants: The semantic integration ensures that Social Determinants of Health (SDoH) factors are explicitly included in the clinical decision-making process, reducing bias and enhancing ethical care delivery.
- Support for Digital Health Infrastructure: The workflow demonstrates the feasibility of using the MMO as a semantic backbone in digital mental health infrastructures, enabling standardized data pipelines for risk assessment, screening, and preventive interventions in psychiatry.

## 3 Discussion

### 3.1 Strengths and Weaknesses

#### Strengths

A technology-driven, interoperable approach in healthcare could offer a number of notable strengths through the Meta Mesh Ontology (MMO), potentially improving care outcomes significantly. Beyond its theoretical foundation, the MMO demonstrates practical impacts on healthcare by enabling the integration of heterogeneous knowledge bases and advancing structured interoperability. By integrating medical, technological, environmental, and socioeconomic data streams through standardized sub-ontologies - including smart devices, sensors, electronic health records, and patient-reported outcomes - a multidimensional picture of patient health emerges, supporting more precise, personalized, and context-sensitive interventions. This structured networking of knowledge bases lays the foundation for optimized precision medicine and preventive strategies based on continuous monitoring of biological, psychological, and environmental influences. Another key advantage of the MMO lies in its interoperability, which could facilitate more efficient use of resources by minimizing redundant tasks, shortening diagnostic times, and optimizing administrative processes. This would be particularly valuable for harmonizing data exchange in globally connected healthcare systems, where the MMO could support international standards and medical terminologies, such as ICD, DSM, SNOMED CT, UMLS, LOINC, DICOM [36], and ISO frameworks to enable interoperable and reliable cross-border information sharing. This alignment ensures semantic interoperability across psychiatry, neuroscience, genetics, and urban health, while also providing a foundation for the responsible development and use of artificial intelligence, consistent with relevant ISO standards and ethical frameworks (see Supplements 12.2). The standardized and structured data management proposed by the MMO provides the foundation for AI-driven data analysis, enabling machine learning approaches to enhance the quality of care through more accurate diagnoses and reduced human error. This capability ensures semantic consistency and interoperability across heterogeneous data sources, thereby establishing the foundation for scalable, explainable, and clinically applicable AI in mental health care. By embedding these standards, the MMO provides not only structural improvements for clinical research and practice but also a future-oriented foundation for ethical, transparent, and socially responsible health technologies. A detailed impact analysis covering these dimensions is provided in the Supplementary Materials. Healthcare providers would thus have access to well-founded, structured, and always up-to-date information, which would contribute to better patient care. In summary, the MMO, as the proposed central component, offers an interoperable, flexible, and scalable framework that could significantly enhance healthcare delivery both on an individual and global scale.

#### Weaknesses

Nevertheless, there are also weaknesses that must be considered. The introduction of interoperable systems is initially cost-intensive and requires extensive adjustments to existing structures. This not only affects the technical implementation but also entails significant training efforts for professionals who will work with the new systems. Another issue is data protection: the increasing integration and networking of personal data heightens the risk of data breaches. In addition to technical security, ethical and legal questions must also be addressed, particularly concerning international data protection standards. Technical complexity presents another challenge. Many existing systems are proprietary and not designed for seamless interoperability. Harmonizing and standardizing these systems requires not only technological adjustments but also institutional and regulatory agreements, which are not always easy to implement. Furthermore, the Meta Mesh Ontology requires continuous maintenance and regular updates to ensure its long-term usability and acceptance. Every technological advancement comes with risks. Therefore, a careful evaluation and targeted management of these challenges are essential to realizing long-term benefits. Implementations in non-WEIRD (western industrialized rich democratic) contexts involve additional challenges, as limited resources, differing regulatory frameworks, and varying digital infrastructures can hinder large-scale adoption. Therefore, implementation strategies should take into account local capacities, socio-technical conditions, and culturally adapted governance models to ensure equitable accessibility and long-term sustainability.

#### 3.2 Implications for Psychiatric Diagnostics and Therapy

The development of the Meta Mesh Ontology (MMO) for psychiatric diagnostics and therapy has far-reaching implications for the interoperability and structuring of clinical data. It enables the standardized integration of heterogeneous information sources, thereby systematically improving diagnostic processes and therapeutic interventions. By integrating semantic concepts, various psychiatric classification systems, taxonomies, and frameworks, such as DSM, ICD, HiTOP, and RDoC, can be harmonized and linked to digital systems. This facilitates a more precise recording and analysis of disease progressions as well as the dynamic adaptation of therapeutic measures based on real-time data. A key advantage of the MMO lies in its ability to integrate smart devices, wearables and other technology-supported systems in an interoperable manner. Wearable technologies and sensors continuously provide psychophysiological data such as heart rate variability, sleep patterns, and activity levels. The ontology ensures that these data are consistently processed and embedded into the clinical context to support diagnoses and create personalized therapy plans. Furthermore, the MMO supports the implementation of intelligent decision-support systems that leverage machine learning and AI-driven analytics to identify risk factors for mental disorders at an early stage. This enables proactive and preventive care by detecting symptoms before they fully manifest. In therapy, MMO-based systems can effectively support digital interventions such as telemedicine and digital therapy applications. Through semantic interoperability, therapy plans can be seamlessly coordinated among different healthcare providers, enhancing both treatment quality and efficiency. Additionally, the ontology contributes to designing health-promoting environments in urban areas. By linking smart city data with psychiatric care models, it becomes possible to analyze environmental factors that influence mental well-being. This allows for the development of evidence-based measures to create health-supportive infrastructures. Overall, the MMO provides a holistic, data-driven foundation for psychiatric diagnostics and therapy. It not only enables more precise and personalized treatment but also considers the societal and ecological factors that are essential for mental health.

## 4 Methods

### 4.1 Mathematical Foundation of the Meta Mesh Ontology

The mathematical framework underlying the Meta Mesh Ontology (MMO) was formalized as an extended property-graph model. This structure provides the computational basis for representing hierarchical, weighted, and infinitely extensible relationships among ontology entities.

### Nodes and Weightings

In this model, the nodes represent data points assigned a weighting that reflects their significance within the graph. Formally, each node can be described as an entity *P*_*i*_, where *i* is an index variable representing the infinite set of nodes:

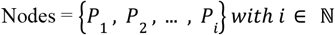

Each node *P*_*i*_ is associated with a weighting ω(*P*_*i*_) that describes its relevance within the context of the data structure:

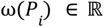

The weighting function provides a formal mechanism for quantifying the relevance or affiliation of ontological entities within the property-graph model. It denotes the proportional assignment of a class or data point to one or multiple knowledge domains and serves both ontological and integrative purposes. On the ontological level, it expresses the degree to which a concept belongs to a given domain; on the integrative level, it quantifies the proportional affiliation of individual data points; on the data level, it describes the evidential strength or frequency within empirical datasets; and on the analytical level, it reflects the contextual importance within a modeling or classification task. All levels follow the same mathematical principle and differ only in their degree of abstraction. Furthermore, each node *P*_*i*_ can possess an infinite number of metadata attributes, which can be described as a function of the form:

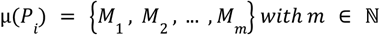

### Domains and Hierarchical Levels

A data point *P*_*i*_ can be organized within one or multiple domains *D*_*i*_. Each domain *D*_*j*_ itself is a node in the graph and is also assigned a weighting:

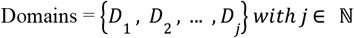

Each domain *D*_*j*_ has a weighting ω(*D*_*j*_), that describes its relevance and can be enriched with an infinite number of metadata attributes:

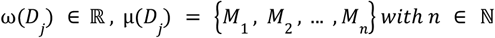

Domains can be further classified into superdomains *S*_*k*_ which assume a higher-level role in the hierarchy. Each superdomain *S*_*k*_ is also a node in the graph with a weighting:

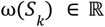

This hierarchical process continues indefinitely, allowing each domain to be embedded within a higher-order superdomain, and each superdomain to be further classified into an even higher level. The hierarchy of domains and superdomains can be described recursively as follows:

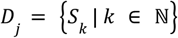

The weighting of a domain *D*_*j*_ can be defined as the sum of the weightings of its higher-level superdomains:

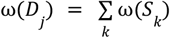

### Edges and Relationships

Edges in the graph represent the relationships between nodes. Each edge connecting a data point *P*_*i*_ to a domain *D*_*j*_ can be defined as:

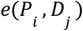

Each edge has an associated weighting that describes the strength or importance of the relationship:

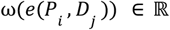

An edge can possess an infinite number of metadata attributes, which can be described as a function:

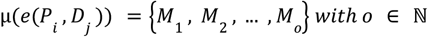

Similarly, edges can also exist between domains and superdomains, denoted as *e*(*D*_*j*_, *S*_*k*_). These edges are also assigned weightings and metadata:

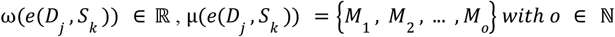

### Recursive Hierarchy and Infinity

The hierarchical structure consists of an infinite sequence of levels, where each node - whether a data point, a domain, or a superdomain - is connected to nodes in the next higher level through edges:

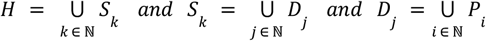

Each entity within this hierarchy, whether a node or an edge, can be enriched with an infinite number of metadata attributes that provide additional contextual information about the respective relationships.

### Overall Hierarchical Structure

In summary, the data structure of the Meta Mesh Ontology (MMO) can be represented as an infinite graph with nested nodes and edges. At each level of the hierarchy, nodes are interconnected by edges, where each edge represents the weight of the relationship and may contain additional metadata. This hierarchy follows a recursive structure and is defined in the form of a hierarchical node *H* as a tuple that encapsulates all relevant information about a data point, domain, or superdomain:

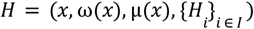

where:

- *x* represents a node (which can be a data point, domain, or superdomain).
- ω(*x*) ∈ ℝ hierarchy. denotes the weighting of the entity, reflecting its relevance in the
- μ(*x*) represents an infinite set of metadata attributes associated with the node.
- {*H*_*i*_}_*i* ∈ *I*_ denotes the set of child nodes hierarchically arranged under xxx, where III is an infinite index set.

Each node *x* can appear at multiple hierarchical levels, and the edges between nodes form a network of relationships that can be analyzed across multiple dimensions. Since both nodes and edges can possess infinite metadata, each connection between entities is not only defined by its weighting but also by additional contextual information stored within the metadata. This hierarchical structure enables the MMO to map heterogeneous knowledge representations and supports multidimensional data analysis, ensuring scalability, interoperability, and adaptability to complex knowledge systems.

### 4.2 Storage and Accessibility

The Meta Mesh Ontology (MMO) was implemented and published using standardized ontology formats, such as OWL (Web Ontology Language) [37] and RDF (Resource Description Framework) [38], which enable structured and formal descriptions of concepts, classes, attributes, relationships, and rules. Persistent and versioned identifiers were established through recognized repositories and version-controlled archives [19,24,39], ensuring long-term reproducibility and traceability. Each ontology element - including classes, properties, and modules - is assigned a unique URI and a persistent URL [40,41], allowing consistent referencing across systems. The ontology is indexed via BioPortal [42,43] and the Ontology Lookup Service (OLS) [44], enabling structured access, semantic search, and cross-ontology linking. Source files, implementation documentation, and example datasets are maintained in the public GitHub repository [45], which also serves as the primary channel for feedback and version tracking [46]. The ontology is distributed under the Creative Commons Attribution 4.0 International (CC BY 4.0) license [47], supporting open reuse and adaptation. This infrastructure ensures that the MMO remains technically accessible, semantically stable, and reproducible for interdisciplinary research and clinical applications.

## 5 Conclusions

The development of the Meta Mesh Ontology (MMO) represents a strategic vision to fundamentally transform psychiatric diagnostics and therapy. Its potential lies not only in the semantic interoperability and standardization of clinical and technological data but also in creating an infrastructure that enables the sustainable and evidence-based advancement of psychiatry. A key aspect of the MMO is the establishment of a methodological foundation that allows the integration of heterogeneous data sources for both scientific and clinical purposes. This involves not only the harmonization of existing classification systems such as DSM, ICD, HiTOP, and RDoC but also the structured incorporation of new insights gained from AI-driven analyses into diagnostic and therapeutic processes. The MMO presents an opportunity to make psychiatric care not only more individualized but also fairer and more sustainable. By incorporating ecological and social factors, it could contribute to the long-term development of resilient healthcare structures and the establishment of evidence-based measures for promoting mental health. In summary, the MMO can be seen as a forward-thinking concept that not only drives technological innovation but also enables a new form of data-driven, interdisciplinary, and ethically informed psychiatry.

## 8 Authors’ contributions

R.B. conceptualized, designed, and developed the complete Meta Mesh Ontology (MMO) and its Core Domain Framework (CDF), performed all analyses, and wrote the manuscript. M.R., A.K., R.L., and S.T. reviewed the manuscript and provided valuable conceptual and structural feedback. N.O. supervised the project, provided critical input on the overall framework, and supported the time allocation and structural guidance for the development and writing of the paper. All authors read and approved the final version of the manuscript.

## Acknowledgements

The author gratefully acknowledges the institutional support from the Jena University Hospital, the Charité-Universitätsmedizin Berlin, and the University of Münster. The author also thanks the German Center for Mental Health (DZPG) for providing the research environment and infrastructure that facilitated the ontology development and semantic validation.

## 10 Data availability

All data necessary to interpret, replicate, and build upon the findings of this study are available. The authoritative ontology artifacts (RDF/XML/OWL) for the Meta Mesh Ontology (MMO) and its Core Domain Framework (CDF) are publicly available in the project’s GitHub repositories. The Supplementary Information provides the complete human-readable definitions of all MMO and CDF classes referenced in the manuscript. Beyond the ontology artifacts, no additional datasets were generated or analyzed for this study.

## 11 Code availability

The code used for ontology generation, validation, and transformation is publicly available in the project’s GitHub repositories. No code is provided with this submission; editors and referees can access the repositories via the links. The repositories contain documentation sufficient to reproduce the results reported in this article.

