## Supplements for "Meta Mesh Ontology: A Transformative Approach to Mental Health Knowledge Integration"

Psychiatric diagnostics struggle to capture the complexity of mental disorders, thereby hampering individualized treatment and personalized psychiatry. Conventional classification systems like DSM-5 and ICD-11 impose rigid categories that inadequately reflect multifactorial, dynamic, and comorbid conditions. To address this limitation, alternative frameworks such as dimensional models and domain ontologies for structured knowledge representation have emerged, yet they often remain isolated and non-interoperable. We present the Meta Mesh Ontology (MMO) as an integrative framework unifying heterogeneous psychiatric knowledge bases through a modular, BFO-compliant structure. The MMO ensures semantic and structural consistency across biological, psychological, and social domains, enabling harmonized data representation and cross-domain reasoning. Its interoperability with established ontologies and informatics standards facilitates integration of multimodal data from clinical, environmental, and sensor sources. By bridging diverse taxonomies such as HiTOP and RDoC, the MMO establishes a scalable foundation for data harmonization and personalized, multidimensional diagnostics in mental health research and practice.

[*] Corresponding author.

[†]

**Keywords:** Meta Mesh Ontology (MMO), Core Domain Framework (CDF), Ontology integration, Semantic interoperability, Clinical data standards, Knowledge representation, Basic Formal Ontology (BFO), Digital medicine infrastructure, FAIR data principles, Biomedical semantics, Data harmonization, Ontology-based clinical research, Interoperable health information systems, Semantic mapping, AI-driven ontology modeling

### 12 Supplements

#### 12.1 Introduction

This supplement provides a comprehensive reference of all classes defined in the Meta Mesh Ontology (MMO) and the Core Domain Framework (CDF). The goal is to document the full semantic landscape of the ontology, ensuring transparency, reproducibility, and interoperability for future research and applications. All class definitions are extracted consistently from the ontology modules, following standardized naming conventions and hierarchical organization.

In addition to the formal ontology structure, this supplement includes a detailed mapping of all data points used in the applied use case (Enabling Integrated Mental Health Care through the Meta Mesh Ontology). This section - Semantic Classification and Psychiatric Relevance of Use Case Data Points - provides a structured table in which each data point is semantically aligned with the MMO/CDF class system. For every data point, the ontological rationale for classification is explained, along with its clinical relevance in psychiatric assessment, diagnosis, or treatment. This mapping demonstrates the practical utility of the ontology in structuring complex, heterogeneous mental health information.

#### 12.2 Analysis of Interoperability and Healthcare Integration

##### 12.2.1 Integration of Heterogeneous Knowledge Bases in Healthcare

To address an ethical, diverse, and personalized psychiatry, the core areas of medicine, technology, socioeconomics, and international standards must be considered with a focus on adaptive and context-based decision-making. The interoperability of such a technology-driven approach in healthcare - integrating smart devices, sensor technology, artificial intelligence, and ISO standards - is enabled through the Meta Mesh Ontology (MMO). This structured networking process is achieved by transforming the knowledge bases of these core areas into sub-ontologies. As a result, the MMO unlocks significant potential for a modern and efficient healthcare system. Heterogeneous systems, platforms, and stakeholders can thus be connected, harmonizing data from various sources such as smart devices, passive sensors, patient-reported outcomes (PROs), and electronic health records (EHRs). This integration enables the creation of more comprehensive and precise patient health profiles, continuous monitoring of vital parameters and environmental influences, and the discovery of novel correlations between previously isolated sources of information. The ability to integrate data from diverse sources - including smart devices, sensors, and electronic health records - into a unified system provides clinical research with access to multidimensional health profiles. These profiles encompass vital signs, patient-reported outcomes, and environmental influences, continuously updated to reflect real-time health status. In summary, the MMO establishes a foundation for an integrative, predictive, and personalized healthcare system that benefits both research and clinical practice. These advancements contribute to improving care quality, optimizing resource utilization, and reducing healthcare costs.

#### 12.2.2 Improving Healthcare Quality Through Structured Integration

The integration of individual health data, including genetic and environmental information, advances precision medicine. A comprehensive and integrated data model enables more precise and context-sensitive study results, supporting the development of personalized therapies. Clinical research can thus design more targeted and well-adapted treatment strategies. A particular focus is placed on identifying factors influencing mental health. Environmental conditions such as noise, air pollution, or social isolation - captured through smart city technologies - can be correlated with a patient's mental and physical health data to detect harmful influences on mental well-being at an early stage. By combining environmental data with patient-reported outcomes, healthcare can be more closely tailored to an individual's specific needs. For example, interventions could be designed to specifically reduce environmental stressors. By merging passively collected environmental data (e.g., air quality, noise levels, social activity) with patient data (e.g., symptoms, well-being, stress levels), a more comprehensive picture of a patient's health status and socioeconomic environment can be drawn. The resulting analyses and preventive measures have the potential to significantly enhance healthcare quality. To ensure these improvements are implemented in a structured and uniform manner, the use of ISO-certified standards provides a common framework for all stakeholders and systems, regardless of manufacturer or technology. These standards help unify data formats, access methods, and security protocols on a global scale, promoting smoother collaboration between different healthcare systems and organizations. This is particularly crucial during global health crises, where resources, data, and expertise must be rapidly shared. An essential aspect of interoperability lies in the more efficient utilization of resources, such as medical personnel and equipment. The optimized data flow enabled by the Meta Mesh Ontology (MMO) reduces redundant efforts and shortens diagnostic times. These efficiency gains not only relieve the burden on medical staff but also lower administrative costs. By seamlessly integrating and harmonizing data from various sources - such as smart devices, sensors, and electronic health records - healthcare processes become more efficient. Moreover, the time dedicated to direct patient care increases due to these streamlined workflows, ultimately leading to improved healthcare quality. For clinical studies and practical healthcare applications, the MMO not only provides structural improvements but also offers potential for innovative therapy approaches and a future-proof healthcare system.

#### 12.2.3 Interoperability with Medical Standards

The Meta Mesh Ontology (MMO) is designed to ensure comprehensive interoperability with existing ontologies, terminologies, classification systems, research frameworks, and knowledge domains in psychiatry. These include SNOMED CT (Systematized Nomenclature of Medicine - Clinical Terms) [32,33], UMLS (Unified Medical Language System) [37,38], and LOINC (Logical Observation Identifiers Names and Codes) [35,36], which enable the semantically precise recording of psychiatric diagnoses, symptoms, treatment approaches, and laboratory data. Additionally, the MMO ensures interoperability with classification systems such as ICD-10/ICD-11 (International Classification of Diseases, 10th/11th Revision) [48-50] and DSM-5 (Diagnostic and Statistical Manual of Mental Disorders, 5th Edition) [51,52]. At the same time, it supports modern, dimensional models of psychopathology, particularly the Hierarchical Taxonomy of Psychopathology (HiTOP) [10,12,53], which describes mental disorders along continuous trait dimensions, and the Research Domain Criteria (RDoC) framework [11,54], which systematizes biological, cognitive, and behavioral markers of psychiatric disorders. A special focus is placed on the integration of neurobiological data. By incorporating the DICOM (Digital Imaging and Communications in Medicine) standard [36], the MMO ensures interoperability with imaging techniques that are increasingly relevant in psychiatric research and diagnostics. These include functional magnetic resonance imaging (fMRI), positron emission tomography (PET), and electroencephalography (EEG). Standardized recording of neural correlates of psychiatric disorders not only optimizes clinical diagnostics but also advances AI-driven analyses of brain activity. Furthermore, the MMO enables the integration of genetic and molecular data, which play a crucial role in personalized psychiatry. Standards such as HGVS (Human Genome Variation Society) [55,56] for genetic variants, OMIM (Online Mendelian Inheritance in Man) [57] for monogenic disorders with psychiatric relevance, and ClinVar [58] for disease-associated genetic variants are incorporated. This facilitates precise annotation of genetic predispositions to psychiatric disorders and promotes the advancement of precision medicine approaches. Another key application area is the standardization of biological sample data. Through interoperability with ISBER (International Society for Biological and Environmental Repositories) [59,60] and MIABIS (Minimum Information About Biobank Data Sharing) [61], structured management of biomaterials such as blood, cerebrospinal fluid, and saliva samples is enabled. By supporting established psychiatric classifications, dimensional models, imaging standards, and genetic data, the MMO provides a central interface for semantic interoperability in psychiatry. It allows for a comprehensive, standardized, and interoperable use of heterogeneous medical and scientific data to further advance diagnostics, research, and therapeutic approaches in psychiatry.

#### 12.2.4 Implementation of International Standards

In this context, the integration of health data with modern technologies such as smart devices, sensors, and IoT systems is essential, offering the potential to incorporate environmental influences and socioeconomic factors into health analysis. By capturing data on environmental conditions, energy efficiency, air quality, and other relevant parameters, a patient’s physical and social surroundings can be comprehensively monitored. This not only enhances the understanding of patients' quality of life but also contributes to the early identification of health risks. However, integrating such data requires the use of international standards that ensure the secure and efficient management of these technologies. The foundation for this is established by standards such as ISO/IEC 30141 for IoT architectures [62], ISO 50001 for energy management [63], ISO/IEC 21451 for smart sensors [64-67], ISO/IEC 20005 for smart cities [68] and ISO/IEC 27000 [69] for data protection regulations. These standards enable the safe and effective use of sensors and smart devices, which capture critical environmental data in real-time and contribute to health analysis. Passive sensors play a central role in this process, as they allow continuous monitoring of relevant environmental conditions without requiring active data input from the patient. By ensuring interoperability through international standards, these technologies can be seamlessly integrated into digital health infrastructures, facilitating more precise, data-driven healthcare solutions.

#### 12.2.5 Health Analysis and Prevention

The collected information - such as air quality, noise levels, and other environmental influences - can be analyzed in real time and compared with individual health profiles. This enables the early identification of potential stressors and the targeted implementation of preventive measures. For example, understanding the correlations between urban environmental factors and health burdens can contribute to long-term improvements in healthcare quality. In urban environments, this data-driven health analysis is increasingly regarded as part of a social ecosystem, where environmental factors, infrastructure, and quality of life are closely interconnected. An interoperable system allows for continuous data collection and analysis, facilitating early detection of health risks and ongoing monitoring. AI-driven algorithms can access this real-time data, improving diagnostic precision and automating the generation of recommendations. Clinical professionals benefit from AI-supported decision-making, as it reduces human errors and enhances patient safety. To analyze environment-related health factors in cities, standardized indicators for urban services and quality of life are required. These include air pollution, noise levels, and traffic density, which influence social and ecological conditions and provide insights into patients' socioeconomic environments. Standards such as ISO 37120 [35], which defines general city indicators, and ISO 37122 [70], which adds specific smart city metrics, form the foundation for such assessments. The collected environmental and social data provide valuable insights for proactive urban planning and healthcare management, fostering healthier living environments and more resilient public health systems.

#### 12.2.6 Artificial Intelligence and Ethics in Healthcare

To effectively analyze and integrate health data into decision-making processes, the use of artificial intelligence (AI) is becoming increasingly important. Standardized frameworks are necessary to ensure the reliable, transparent, and ethically responsible use of AI in healthcare. A unified terminology and methodological guidelines facilitate the seamless integration of AI-powered systems within medical environments. Key considerations include guidelines for transparency, ethical principles, and risk management, as well as measures to prevent algorithmic discrimination and protect sensitive health data. These aspects are addressed through various standards, including ISO/IEC 22989 (Terminology) [71], ISO/IEC 23053 (Machine Learning Framework) [72], ISO/IEC 23894 (Risk Management) [73], ISO/IEC 24027 (Bias Reduction) [74], ISO/IEC 29100 (Data Protection) [75] and the technical recommendation ISO/IEC TR 24028 [76] for AI trustworthiness. While technical standards provide the foundation for the secure and efficient deployment of AI, they alone are insufficient to ensure a sustainable and human-centered approach. Ethical considerations, regulatory oversight, and continuous evaluation of AI systems are essential to maintaining trust and fairness in AI-driven healthcare applications.

#### 12.2.7 Ethical and Social Responsibility

Ethical, social, and work-related aspects are equally crucial, especially when AI is implemented in sensitive fields such as healthcare. Therefore, the integration of social, ethical, and organizational standards is recommended. Social responsibility, diversity and inclusion management, and a human-centered organizational structure contribute to upholding patient dignity. Moreover, workplace mental health and the importance of fairness and ethical conduct in business processes are of particular relevance. The corresponding guidelines can be found in standards such as ISO 26000 (Social Responsibility) [77], ISO 30415 (Diversity and Inclusion) [78], ISO 27500 (Human-Centered Organization) [79], ISO 45003 (Psychological Health) [80] and ISO 37001 (Ethical Conduct) [81]. Integrating ISO standards for smart cities, sensor technology, and IoT into a health monitoring system could provide an enhanced, data-driven perspective on patients' social and economic environments. This approach could be highly beneficial for both the prevention and treatment of mental and physical illnesses. By combining passive sensor data with patient-reported outcomes, a comprehensive view of patients and their surroundings can be achieved, ultimately leading to better, more personalized healthcare. By applying these responsible, transparent, and fair standards, ethical concerns are addressed, data privacy is safeguarded, and health technology is ensured to serve the well-being of both patients and society.

### 12.3 Method for Deriving the Core Domain Framework (CDF) Domains

To ensure maximum cross-domain interoperability and comprehensive knowledge integration within the Meta Mesh Ontology (MMO), the Core Domain Framework (CDF) was developed as the top-level abstraction layer. The purpose of this framework is to provide a consistent domain scaffold aligned with the MMO, under which heterogeneous domain-specific MMO modules can be integrated - without semantic redundancy or fragmentation.

The derivation of the CDF domains followed a multi-stage conceptual synthesis guided by both epistemological principles and semantic interoperability requirements. Philosophical-ontological analyses, crosswalks between existing domain vocabularies, and pragmatic modeling needs in the context of personalized psychiatry and biomedical knowledge integration were combined.

#### 12.3.1 Ontological Anchoring

The starting point was the ontological structure of the Meta Mesh Ontology (MMO), which, as its top-level ontology, uses the Basic Formal Ontology (BFO) and distinguishes between continuants (e.g., dispositions, qualities, independent entities) and occurrents (e.g., processes). The CDF is designed to function as an intermediary domain layer within this structure. Each CDF class is ontologically designed to serve as a superclass for domain-specific MMO modules and to align with an appropriate MMO category.

#### 12.3.2 Philosophical and Epistemic Grounding

Philosophical models of knowledge organization were reviewed, particularly those attempting to partition human knowledge into exhaustive and mutually exclusive domains. Relevant contributions included Aristotle’s theoretical-practical-productive classification, the DDC and LCC taxonomies, and modern philosophical reconstructions of scientific domains.

#### 12.3.3 Analysis of International Classification Systems

To ensure the CDF domains are compatible with international standards and domain vocabularies, a selection of global classification and semantic systems was analyzed:

- UNESCO Thesaurus (for disciplinary breadth and ontology-compatibility)
- OECD Fields of Science (FOS) (for alignment with science systems)
- ISO 11179 (for structured metadata modeling)
- DDC / LCC (for validated hierarchical knowledge coverage)
- Wikidata / DBpedia (for web interoperability)
- UN SDGs (for policy and sustainability contexts)

By triangulating these sources, recurring high-level distinctions were identified and their semantic cores mapped to emerging domain patterns in the MMO use cases.

#### 12.3.4 Identification and Abstraction of Domains

From this conceptual synthesis, a compact and abstract set of eight universal domains was derived, each covering a disjoint, semantically coherent, and ontologically generalizable area of human knowledge. These eight CDF top-level domains form the integrative backbone for all subordinate MMO modules:

- **Knowledge**: The domain of knowledge, science, education, and knowledge production - including formal theories, models, and information systems.
- **Culture**: The domain of cultural expressions, values, symbol systems, identities, and worldviews.
- **Health**: The domain of health, disease, prevention, care, treatment, and medical knowledge.
- **Nature**: The domain of the natural world, including biological, physical, chemical, and ecological systems.
- **Technology**: The domain of human-made systems, tools, artifacts, information and communication technologies, and engineering constructs.
- **Economy**: The domain of economic activities, resource allocation, markets, production, exchange, and value creation.
- **Governance**: The domain of normative, legal, political, and institutional steering as well as collective decision-making structures.
- **Society**: The domain of social relations, social structures, roles, communities, and interactions in human coexistence.

These domains were chosen to be non-overlapping while remaining flexible through subclassing.

#### 12.3.5 Semantic Linkage via SKOS

While the CDF classes were modeled as OWL Class constructs to allow full logical reasoning and subclassing, SKOS properties (e.g., skos:exactMatch, skos:closeMatch, skos:broadMatch) were additionally applied to link them to equivalent or related concepts in reference systems. This enables external alignment without compromising the internal logical structure of the ontology.

#### 12.3.6 Conclusion

The resulting CDF provides a domain-agnostic, MMO- and BFO-compliant classification scaffold for modular ontology integration. It ensures semantic clarity, reduces redundancy, and offers a robust basis for systematic cross-domain modeling - particularly suited to the complexities of personalized psychiatry, where biomedical, psychological, environmental, social, and technological dimensions must be represented in an integrated manner.

### 12.4 Ontology Classes

All MMO and CDF class definitions follow the Two-Part Definition Rule, expressed in the format:

**S = Def. a G that D.**

- **S** = the class label to be defined (exact label from the ontology).

- **G** = the immediate parent class in the ontology, expressed as a clear and ontologically robust English term (e.g., *process*, *disposition*, *specifically dependent continuant*).

- **D** = the differentia, i.e., the distinguishing characteristic(s) that make some G's into instances of S. This must be stated precisely, without circularity, unnecessary jargon, or examples.

Each definition is written as a concise, human-readable sentence. The structure ensures that every class is situated within the broader ontology while being differentiated in an ontologically rigorous way. All MMO classes follow the wording and naming conventions of the Basic Formal Ontology (BFO), whereas all CDF classes are represented in concatenated CamelCase notation. This representation was chosen to reflect the distinction between ontology layers and the formal naming conventions within the RDF/XML structure.

### 12.4.1 Meta Mesh Ontology Classes

| Class | Definition |
|---|---|
| entity unit | Def. an entity that exists as an identifiable, delimited unit within a broader context. |
| continuant unit | Def. a continuant that exists as an identifiable, delimited unit within a broader context while maintaining persistence and identity over time. |
| generically dependent continuant unit | Def. a generically dependent continuant that exists as an identifiable, delimited unit instantiated in one or more independent continuants while maintaining its type identity across these instances. |
| independent continuant unit | Def. an independent continuant that exists as an identifiable,delimited unit while maintaining its type identity and spatial location across time. |
| immaterial entity unit | Def. an immaterial entity that exists as an identifiable,delimited unit in relation to material entities and that can undergo change as its material hosts undergo change. |
| continuant fiat boundary unit | Def. a continuant fiat boundary that exists as an identifiable,delimited unit in relation to a material or immaterial entity and that persists while maintaining its identity across changes in the entity it bounds. |
| zero-dimensional continuant fiat boundary unit | Def. a zero-dimensional continuant fiat boundary that exists as an identifiable,delimited unit in relation to a material or immaterial entity and that persists while maintaining its identity across changes in the entity it bounds. |
| one-dimensional continuant fiat boundary unit | Def. a one-dimensional continuant fiat boundary that exists as an identifiable,delimited unit in relation to a material or immaterial entity and that persists while maintaining its identity across changes in the entity it bounds. |
| two-dimensional continuant fiat boundary unit | Def. a two-dimensional continuant fiat boundary that exists as an identifiable,delimited unit in relation to a material or immaterial entity and that persists while maintaining its identity across changes in the entity it bounds. |
| site unit | Def. a site that exists as an identifiable, delimited unit in relation to a material entity and persists while maintaining its identity across changes in the entity that bounds it. |
| spatial region unit | Def. a spatial region that exists as an identifiable, delimited unit within a frame of reference while maintaining its identity across changes in its relational context. |
| zero-dimensional spatial region unit | Def. a zero-dimensional spatial region that exists as an identifiable,delimited unit occupying a single point within a spatial frame of reference. |
| one-dimensional spatial region unit | Def. a one-dimensional spatial region that exists as an identifiable, delimited unit extending along a single dimension within a spatial frame of reference. |
| two-dimensional spatial region unit | Def. a two-dimensional spatial region that exists as an identifiable, delimited unit extending along two dimensions within a spatial frame of reference. |
| three-dimensional spatial region unit | Def. a three-dimensional spatial region that exists as an identifiable, delimited unit extending along three dimensions within a spatial frame of reference. |

| | |
|---|---|
| material entity unit | Def. a material entity that exists as an identifiable, delimited unit composed of some portion of matter while maintaining its type identity across changes in its material composition. |
| fiat object part unit | Def. a fiat object part that exists as an identifiable, delimited material entity that is, at all times of its existence, a proper continuant part of some object and demarcated from the remainder of that object by a two-dimensional continuant fiat boundary. |
| object unit | Def. an object that exists as an identifiable, delimited material entity manifesting maximal causal unity among its material parts under typical conditions for its type. |
| object aggregate unit | Def. an object aggregate that exists as an identifiable, delimited material entity composed of a plurality of objects as member parts while maintaining its identity across changes in membership. |
| specifically dependent continuant unit | Def. a specifically dependent continuant that exists as an identifiable, delimited dependent entity that is realized in or manifested on one or more independent continuants while maintaining its type identity across these instances. |
| quality unit | Def. a quality that exists as an identifiable, delimited dependent entity that is manifested in an independent continuant without requiring any further process for its realization. |
| relational quality unit | Def. a relational quality that is manifested as an identifiable, delimited dependent entity inhering in two or more independent continuants simultaneously. |
| realizable entity unit | Def. a realizable entity that is manifested as an identifiable, delimited dependent entity inhering in an independent continuant and capable of being realized in processes of a correlated type. |
| disposition unit | Def. a disposition that is manifested as an identifiable, delimited dependent entity inhering in a material entity and is realized in virtue of the bearer’s physical constitution under specific conditions. |
| function unit | Def. a function that is manifested as an identifiable, delimited dependent entity inhering in a material entity and is realized in processes of a correlated type in virtue of the bearer’s physical constitution, which exists due to evolutionary development or intentional design. |
| role unit | Def. a role that is manifested as an identifiable, delimited dependent entity inhering in a bearer situated in specific physical, social, or institutional circumstances, and that is realized in correlated processes without requiring a change in the bearer’s physical constitution. |
| occurrent unit | Def. an occurrent that is manifested as an identifiable, delimited entity unfolding in time or existing as the instantaneous boundary of such unfolding, and that occupies a spatiotemporal region corresponding to its temporal extension. |
| process unit | Def. a process that is manifested as an identifiable, delimited occurrent unfolding in time, specifically depending on a material entity, and having proper temporal parts corresponding to its temporal extension. |

| | |
|---|---|
| history unit | Def. a history that is manifested as the total process occurring in the spatiotemporal region occupied by a material entity or site, including all surface and internal processes for which it serves as host. |
| process profile unit | Def. a process profile that is manifested as a proper occurrent part of a process with which it shares a temporal region and is mutually dependent on a distinct, non-overlapping occurrent part of that process. |
| process boundary unit | Def. a process boundary that is manifested as a temporal part of a process and occupies a zero-dimensional temporal region. |
| spatiotemporal region unit | Def. a spatiotemporal region that is manifested as an occurrent occupying a region of spacetime and projecting onto corresponding spatial and temporal regions. |
| temporal region unit | Def. a temporal region that is manifested as an occurrent occupying a portion of time within a reference frame and whose parts are also temporal regions. |
| zero-dimensional temporal region unit | Def. a temporal region that is without temporal extension and occupies a single instant. |
| one-dimensional temporal region unit | Def. a temporal region that is extended along a single dimension of time without gaps or breaks. |

**Table 2.** *An overview of the core ontology classes defined within the Meta Mesh Ontology (MMO). The first column lists the class name as used in the ontology, while the second column contains its formal definition or intended semantic scope.*

### 12.4.2 Core Domain Framework Classes

| Class | Definition |
|---|---|
| KnowledgeBearingEntity | Def. an independent continuant unit that embodies epistemic states within a domain-specific layer without requiring material constitution. |
| CulturalBearingEntity | Def. an independent continuant unit that embodies domain-specific cultural entities and enables the manifestation of cultural functions and artifacts. |
| HealthBearingEntity | Def. an independent continuant unit that embodies domain-specific health-related entities providing for the structured representation of anatomical or clinical physical entities. |
| NaturalBearingEntity | Def. an independent continuant unit that embodies domain-specific natural entities and provides a structured representation of natural physical entities. |
| TechnicalBearingEntity | Def. an independent continuant unit that embodies domain-specific technical entities and provides a structured representation layer for technical physical entities. |

| | |
|---|---|
| EconomicBearingEntity | Def. an independent continuant unit that embodies domain-specific economic entities to enable structured integration of economically relevant physical entities and their process interactions. |
| GovernanceBearingEntity | Def. an independent continuant unit that embodies domain-specific governance-relevant entities enabling the structured representation of politically and administratively significant physical entities. |
| SocialBearingEntity | Def. an independent continuant unit that embodies domain-specific socially relevant entities and functions as a bearer of social roles and relations. |
| NaturalRegion | Def. an immaterial entity unit that embodies domain-specific natural phenomena as spatially extended but non-material entities within ecological or geophysical contexts. |
| SymbolicFiatBoundary | Def. a fiat boundary unit that establishes spatial distinctions based on symbolic or historical criteria without material demarcation. |
| AnatomicalFiatBoundary | Def. a fiat boundary unit that establishes non-material demarcations within the body for precise anatomical localization and the planning of clinical interventions. |
| TechnicalSystemBoundary | Def. a fiat boundary unit that establishes non-material demarcations within technical systems for the structured modeling of constructive relationships. |
| EcologicalBoundary | Def. a fiat boundary unit that establishes non-material demarcations of ecological zones for the scientific classification of environmental areas. |
| JurisdictionalBoundary | Def. a fiat boundary unit that determines non-material demarcations of jurisdictions for the legal and administrative definition of areas of responsibility. |
| SocialBoundary | Def. a fiat boundary unit that establishes non-material demarcations within societies for determining social areas of responsibility and influence. |
| EpistemicSite | Def. a site unit that serves as a non-material domain for the generation, analysis, or dissemination of knowledge. |
| CulturalSite | Def. a site unit that serves as a non-material domain for the enactment and preservation of cultural meanings and practices. |
| ClinicalSite | Def. a site unit that serves as a non-material domain for medical examinations and treatments. |
| NaturalSite | Def. a site unit that is defined as a non-material domain by ecological, geological, or climatological characteristics. |

| | |
|---|---|
| TechnicalSite | Def. a site unit that is defined as a non-material domain by technical functions within a constructed system. |
| EconomicSite | Def. a site unit that is defined as a non-material domain by economic functions and uses. |
| SocialSite | Def. a site unit that serves as a non-material domain for the enactment and structuring of social interactions. |
| CulturalSpatialRegion | Def. a spatial region unit that is defined as a non-material domain by cultural meanings and symbolic relations. |
| AnatomicalSpatialRegion | Def. a spatial region unit that is used as an abstract domain within the body for clinical, anatomical, or diagnostic description. |
| GeoSpatialRegion | Def. a spatial region unit that is used as an abstract domain for the delineation of geophysical or ecological conditions. |
| TechnicalSpatialRegion | Def. a spatial region unit that is used as an abstract domain to support technical design, modeling, and operational processes. |
| EconomicSpatialRegion | Def. a spatial region unit that is used as an abstract domain for structuring economic activities and infrastructures. |
| GovernanceSpatialRegion | Def. a spatial region unit that is used as an abstract domain for delineating areas of responsibility and administration within the context of political and administrative governance. |
| SocialSpatialRegion | Def. a spatial region unit that is used as an abstract domain for localizing and structuring social relations within societies. |
| CulturalPhysicalEntity | Def. a material entity unit that embodies domain-specific cultural entities as delineated physical carriers of cultural functions and expressions. |
| HealthPhysicalEntity | Def. a material entity unit that embodies domain-specific health-related entities as identifiable physical structures relevant for medical and clinical contexts. |
| NaturalPhysicalEntity | Def. a material entity unit that embodies domain-specific natural entities as discrete,independently existing material continua within the realm of nature. |
| TechnicalPhysicalEntity | Def. a material entity unit that embodies domain-specific technological entities as discrete, independently existing material units within the technological domain. |
| EconomicPhysicalEntity | Def. a material entity unit that embodies domain-specific economic entities as discrete, independently existing material units within the economic domain. |

| | |
|---|---|
| GovernancePhysicalEntity | Def. a material entity unit that embodies domain-specific governance-related entities as discrete, independently existing material units within the realm of governance. |
| SocietalPhysicalEntity | Def. a material entity unit that embodies domain-specific societal entities as discrete, independently existing material units within the societal domain. |
| SymbolicFiatPart | Def. a fiat object part unit that exists as a symbolically or functionally delineated component of a cultural object. |
| AnatomicalFiatSegment | Def. a fiat object part unit that exists as a non-naturally delineated section of an organ or body area used for medical description and planning. |
| NaturalFiatPart | Def. a fiat object part unit that exists as a conventionally delineated section of a natural system used for scientific description and analysis. |
| TechnicalFiatPart | Def. a fiat object part unit that exists as a functionally delineated component of a technical system used for the structuring and analysis of technical units. |
| AdministrativeDivision | Def. a fiat object part unit that exists as a conventionally delineated part of a governmental or organizational system used for political or administrative structuring. |
| EconomicTerritorialSegment | Def. a fiat object part unit that exists as an economically relevant, conventionally delineated part of a geographic or administrative territory used for the governance of economic matters. |
| SocialBoundaryRegion | Def. a fiat object part unit that exists as a socially defined, conventionally delineated area used to mark belonging or separation within social groups. |
| CulturalArtifact | Def. an object unit that, as a material entity within a cultural context, embodies symbolic or semantic meanings. |
| AnatomicalStructure | Def. an object unit that, as a material component of a body, possesses a distinct physical identity. |
| NaturalObject | Def. an object unit that exists as a material component of the natural environment. |
| TechnicalDevice | Def. an object unit that is used as a functional-technological unit for carrying out technical. |
| EconomicGood | Def. an object unit that is used as a physical, tradable item in economic transactions. |

| | |
|---|---|
| GovernanceInstrument | Def. an object unit that is used as a material bearer of legal or political authority for the implementation, enforcement, or symbolization of regulatory and administrative functions. |
| CulturalArtifactAggregate | Def. an object aggregate unit that is used as a culturally significant grouping of multiple material items into a single unit. |
| AnatomicalAggregate | Def. an object aggregate unit that consists of multiple biological parts as a functional unit in the anatomical context. |
| NaturalAggregate | Def. an object aggregate unit that consists of multiple natural objects forming a functional or semantic unit. |
| TechnicalModuleAggregate | Def. an object aggregate unit that consists of multiple technical systems or modules that are operated together or managed as a unit. |
| CommodityBatch | Def. an object aggregate unit that encompasses an economically relevant quantity of physical goods as a unit. |
| AdministrativeUnitAggregate | Def. an object aggregate unit that encompasses a group of persons delineated by political or administrative affiliation as a unit. |
| SocialGroupAggregate | Def. an object aggregate unit that embodies a unit composed of individual persons through social affiliation, shared activities, or shared identity. |
| EpistemicState | Def. a specifically dependent continuant unit that realizes knowledge as cognitive states within individual bearer systems. |
| CulturalCondition | Def. a specifically dependent continuant unit that realizes cultural meanings or roles within individual bearer systems. |
| ClinicalState | Def. a specifically dependent continuant unit that realizes health conditions or disease courses within individual bearers. |
| NaturalPropertyState | Def. a specifically dependent continuant unit that realizes physical or biological states within individual natural objects. |
| OperationalStatus | Def. a specifically dependent continuant unit that realizes the operational state of an individual technical system during a specific temporal phase. |
| EconomicCondition | Def. a specifically dependent continuant unit that embodies the economic condition of an individual actor or system during a specific phase. |
| GovernanceCondition | Def. a specifically dependent continuant unit that embodies an institutionally bound condition of a political actor or authority within a specific governance-related context. |

| | |
|---|---|
| SocialCondition | Def. a specifically dependent continuant unit that embodies a social status or social role of an individual within a specific social context. |
| EpistemicQuality | Def. a quality unit that provides intrinsic properties of epistemic entities as characteristics describing the certainty, plausibility, accuracy, or reliability of knowledge content or conceptual structures. |
| SymbolicQuality | Def. a quality unit that provides perceivable, symbolic features of a cultural object or cultural practice in the form of stylistic, ornamental, or motif expressions. |
| PhysiologicalQuality | Def. a quality unit that provides physical or biological characteristics of a living organism or its parts in the form of measurable states. |
| NaturalPhenotypeQuality | Def. a quality unit that provides intrinsic physical or biological characteristics of a natural entity as perceptible properties. |
| TechnicalPerformanceQuality | Def. a quality unit that provides measurable properties of technical artifacts or systems for describing and evaluating technical performance. |
| EconomicValueQuality | Def. a quality unit that provides assessable economic characteristics of actors or systems within a broader economic framework. |
| GovernanceResponsivenessQuality | Def. a quality unit that provides qualitative characteristics of governance or administrative processes as assessable properties within governance evaluation. |
| SocialCohesionQuality | Def. a quality unit that describes characteristics expressing cohesion, trust, and integration within a social group or community. |
| EpistemicRelationalQuality | Def. a relational quality unit that provides comparative or relational aspects between epistemic entities for describing dependencies, contrasts, or inferential structures. |
| SymbolicRelationalQuality | Def. a relational quality unit that provides culturally meaningful relational properties as structured relations between cultural actors or artifacts for describing symbolic contrasts or roles. |
| PhysiologicalRelationalQuality | Def. a relational quality unit that provides qualitative relations between physiological entities or parameters for describing asymmetries, gradients, or comparative measurements in clinical and health-related contexts. |
| EcologicalRelationalQuality | Def. a relational quality unit that describes qualitative characteristics realized as relations between organisms or |

| | |
|---|---|
| | environmental factors within ecosystems in the form of interactions such as symbiosis, competition, or spatial distribution. |
| TechnicalConnectivityQuality | Def. a relational quality unit that provides qualitative characteristics realized as connections and interactions between components of technical systems in the form of compatibility, signal integrity, or latency. |
| EconomicRelationalState | Def. a relational quality unit that realizes economic relations such as trade imbalances, investment ratios, or hierarchical governance structures between economic actors or institutions as relational states within their interactions. |
| GovernanceRelationalQuality | Def. a relational quality unit that realizes power, responsibility, and authority relations within political or administrative structures as dependencies between institutional actors within their interactions. |
| SocialRelationalQuality | Def. a relational quality unit that realizes interpersonal or group-related relations as dependencies between actors within social interactions. |
| KnowledgeCapacity | Def. a realizable entity unit that provides for the potential realization of cognitive acts and states enabling recognition, hypothesis formation, and theory development. |
| CulturalCapacity | Def. a realizable entity unit that provides for the realization of participation in symbolic practices and meaning-making within ritual and interpretive acts. |
| HealthCapability | Def. a realizable entity unit that provides for the realization of self-regulation, immune response, and healing potentials within an organism. |
| NaturalPotential | Def. a realizable entity unit that provides for the realization of potentials inherent in natural phenomena, such as growth, photosynthesis, or erosion. |
| TechnicalCapacity | Def. a realizable entity unit that provides for the realization of functional capacities in machines, software, and systems within technical operations. |
| EconomicCapacity | Def. a realizable entity unit that is manifested as capacities for payment, tradability, or liquidity within economic interactions. |
| GovernanceMandate | Def. a realizable entity unit that is manifested as an authorization or capacity for rule-setting, authorization, or decision-making within governance processes. |

| | |
|---|---|
| SocialCompetence | Def. a realizable entity unit that is manifested as the capacity for role assumption, social interaction, and adherence to norms within societal structures. |
| EpistemicDisposition | Def. a disposition unit that realizes the tendency to generate, convey, or maintain specific interpretations or meanings as a culture- or symbol-bound property in an actor or artifact. |
| PersonCenteredDisposition | Def. a disposition unit that is realized as a tendency anchored in an individual person toward health-related states, functions, or behaviors. |
| ClinicalDisposition | Def. a PersonCenteredDisposition that provides the potential for the onset, course, or modification of a pathological or health-relevant state in a biological organism under specific biological or environmental conditions. |
| PsychologicalDisposition | Def. a PersonCenteredDisposition that provides the tendency to manifest specific mental states or behavior patterns in a person in response to psychological or social stimuli. |
| NaturalDisposition | Def. a disposition unit that provides the tendency to realize specific physical or biological reactions in a natural object under appropriate environmental conditions. |
| TechnicalDisposition | Def. a disposition unit that provides the tendency to perform or withhold technical functions in an artifact or technical system under specific physical or operational conditions. |
| EconomicDisposition | Def. a disposition unit that provides the tendency of an economic actor or system to respond to financial or market-related conditions under specific economic situations. |
| SocialDisposition | Def. a disposition unit that provides the tendency of an individual or group to manifest certain social behaviors or attitudes in interpersonal or group-related interactions. |
| EpistemicFunction | Def. a function unit that provides the role or capacity for generating, disseminating, or verifying knowledge in an actor or system. |
| SymbolicFunction | Def. a function unit that provides the role for fulfilling symbolic purposes in a cultural artifact or practice. |
| PhysiologicalFunction | Def. a function unit that provides the role for performing biological processes in a body part under physiological conditions. |
| EcologicalFunction | Def. a function unit that provides the role in contributing to ecological processes by natural entities within environmental contexts. |

| | |
|---|---|
| TechnicalFunction | Def. a function unit that enables the performance of specific technical tasks by intentionally designed artifacts or systems to provide planned effects. |
| EpistemicRole | Def. a role unit that provides the situational involvement of an actor in processes of knowledge generation or validation within institutional or discursive frameworks. |
| CulturalRole | Def. a role unit that provides the situational involvement of a person or object in cultural practices as part of social processes. |
| ClinicalRole | Def. a role unit that provides the institutionally and socially assigned participation of a person in medical processes within a clinical context. |
| NaturalResourceRole | Def. a role unit that provides the environmental use, management, or intended purpose of a natural resource within ecological or governance-related processes. |
| TechnicalRole | Def. a role unit that is assigned to a technical artifact to fulfill a specific, situational function within a technical system. |
| EconomicRole | Def. a role unit that is assigned to a person or organization to fulfill a specific function within an economic system in a context-dependent situation. |
| GovernanceRole | Def. a role unit that is assigned to a person within a political or administrative system for carrying out specific tasks and authorities in institutional contexts. |
| SocialRole | Def. a role unit that is assigned to a person within social contexts for fulfilling certain tasks, expectations, or relationships through social interactions and normative structures. |
| EpistemicProcess | Def. a process unit that is realized as a temporally extended activity for the generation, verification, or dissemination of knowledge involving actors and media. |
| CulturalProcess | Def. a process unit that is realized as a temporally extended sequence of actions with cultural significance involving actors and materials. |
| HealthProcess | Def. a process unit that is realized in the emergence, maintenance, transformation, or observation of states in living organisms within a health-related context. |
| ClinicalProcess | Def. a process unit that is realized as a temporally extended sequence of actions or events for diagnosis, treatment, or observation of health conditions within a clinical context. |
| PhysiologicalProcess | Def. a HealthProcess that is realized in internal bodily functions of an organism without intentional control. |

| | |
|---|---|
| NaturalProcess | Def. a process unit that is realized as a temporally extended sequence of events or state changes in natural systems without intentional control. |
| TechnicalProcess | Def. a process unit that is realized as a temporally extended, functionally intended sequence of operations in technical systems for achieving specific technical purposes. |
| EconomicProcess | Def. a process unit that is realized as a temporally extended sequence of economic activities for the production, distribution, or use of goods and services within economic systems. |
| GovernanceProcess | Def. a process unit that is realized as an institutionally structured and temporally bounded sequence of actions for regulation, control, or decision-making within political or administrative systems. |
| SocialProcess | Def. a process unit that is realized as a temporally structured and interactive sequence of actions between social actors for shaping social relationships or collective actions. |
| EpistemicDevelopmentHistory | Def. a history unit that is realized as a temporally structured sequence of cognitive processes, social interactions, and documented artifacts for representing the development of knowledge. |
| CulturalHistory | Def. a history unit that is realized as a temporally structured sequence of cultural practices, social interactions, and material artifacts for representing the development of cultural phenomena. |
| ClinicalHistory | Def. a history unit that is realized as a temporally structured sequence of medically relevant events and states for representing the course of health and disease processes in an individual. |
| EcologicalHistory | Def. a history unit that is realized as a temporally structured sequence of natural processes and states for representing the development of ecosystems, landscapes, or species. |
| TechnologicalHistory | Def. a history unit that is realized as a temporally structured sequence of processes and states for representing the origin and development of technical systems. |
| EconomicHistory | Def. a history unit that is realized as a temporally structured sequence of processes and states for representing the development of economic activities and structures. |
| GovernanceHistory | Def. a history unit that is realized as a temporally structured sequence of institutional, legal, and social processes for representing political developments. |

| | |
|---|---|
| SocialHistory | Def. a history unit that is realized as a temporally structured sequence of role changes, interaction dynamics, and collective experiences for representing societal changes. |
| EpistemicProcessProfile | Def. a process profile unit that is realized as a structured temporal development in cognitive or knowledge-generating processes for describing changes in insight, uncertainty, or coherence. |
| CulturalExpressionProfile | Def. a process profile unit that is realized as a structured temporal and formal manifestation of cultural performances, rituals, or collective narratives in cultural processes. |
| DiseaseProgressionProfile | Def. a process profile unit that is realized as a characteristic temporal manifestation of disease progression phases within disease-related processes. |
| NaturalDynamicsProfile | Def. a process profile unit that is realized as a structured temporal manifestation of patterns in natural processes for describing dynamics in ecological or geological changes. |
| TechnicalOperationProfile | Def. a process profile unit that is realized as a structured temporal manifestation of behavior patterns during the functioning of technical systems for describing system dynamics and operational characteristics. |
| MarketFluctuationProfile | Def. a process profile unit that is realized as a structured temporal manifestation of fluctuation patterns in economic processes for describing dynamic market developments. |
| GovernanceDecisionProfile | Def. a process profile unit that is realized as a structured temporal manifestation of decision-making sequences in institutional or regulatory processes for describing dynamics of political and legal decision-making. |
| SocialInteractionProfile | Def. a process profile unit that is realized as a structured temporal manifestation of social dynamics in interaction processes for describing patterns of social sequences. |
| EpistemicTransitionPoint | Def. a process boundary unit that is realized as a sharply delineated point in time of change in knowledge-generating processes to mark the transition between epistemic phases. |
| CulturalTransitionPoint | Def. a process boundary unit that is realized as a temporal marker of the beginning or end of cultural actions in normative or symbolically influenced contexts. |
| ClinicalTransitionPoint | Def. a process boundary unit that is realized as a temporally clearly delineated transition within medical processes for the recognition, monitoring, and documentation of clinical events. |

| | |
|---|---|
| NaturalTransitionPoint | Def. a process boundary unit that is realized as a temporally clearly determinable transition within natural processes for delineating phases in ecological or geophysical systems. |
| TechnicalTransitionPoint | Def. a process boundary unit that is realized as a temporally determined transition between operational states of technical systems for delineating functional phases. |
| EconomicTransitionPoint | Def. a process boundary unit that is realized as a temporally determined transition between states of economic processes for delineating economic dynamics. |
| PoliticalTransitionPoint | Def. a process boundary unit that is realized as a temporally determined transition between states of political processes for delineating institutional changes. |
| SocialTransitionPoint | Def. a process boundary unit that is realized as a temporally determined transition between states of social processes for delineating social interactions. |
| EpistemicSpacetimeContext | Def. a spatiotemporal region unit that is realized as the spatial-temporal framework for the execution, mediation, or reception of knowledge-related processes. |
| CulturalSpacetimeContext | Def. a spatiotemporal region unit that is realized as the spatial-temporal framework for the enactment of cultural expression processes. |
| ClinicalSpacetimeContext | Def. a spatiotemporal region unit that is realized as the spatial-temporal framework for the conduct of medical interactions. |
| NaturalSpacetimeContext | Def. a spatiotemporal region unit that is realized as the spatial-temporal framework for the occurrence of natural processes. |
| TechnicalSpacetimeContext | Def. a spatiotemporal region unit that is realized as the spatial-temporal framework for the occurrence of technical operations. |
| EconomicSpacetimeContext | Def. a spatiotemporal region unit that is realized as the spatial-temporal framework for the occurrence of economic processes. |
| GovernanceSpacetimeContext | Def. a spatiotemporal region unit that is realized as the spatial-temporal framework for the occurrence of governance activities and political decision-making processes. |
| SocialSpacetimeContext | Def. a spatiotemporal region unit that is realized as the spatial-temporal framework for the occurrence of social events and interactions. |

| | |
|---|---|
| EpistemicTimeFrame | Def. a temporal region unit that is realized as the time frame for delineating phases of epistemic relevance in knowledge development. |
| CulturalTimeFrame | Def. a temporal region unit that is realized as the time frame for delineating epochs with defining cultural characteristics. |
| TherapeuticWindow | Def. a temporal region unit that is realized as the time frame for the execution or effectiveness of therapeutic interventions. |
| EcologicalTimeFrame | Def. a temporal region unit that is realized as the time frame for the occurrence and course of ecological processes. |
| TechnicalTimeFrame | Def. a temporal region unit that is realized as the planned or measured period for technical processes and states. |
| EconomicTimeFrame | Def. a temporal region unit that is realized as a delineated period for the analysis and structuring of economic processes and cycles. |
| GovernanceTimeFrame | Def. a temporal region unit that is realized as a formally defined period for structuring and analyzing legal and institutional governance processes. |
| SocialTimeFrame | Def. a temporal region unit that is realized as an abstract period for structuring and analyzing social development processes. |

**Table 3.** *An overview of the foundational ontology classes defined within the Core Domain Framework (CDF) of the Meta Mesh Ontology (MMO). The first column lists the class name, while the second column provides a formal definition describing its intended scope and role within the ontology.*

### 12.5 Domain-Aligned Representation of Heterogeneous Data Points

| Domain | MMO Class | Datapoint | Code | Code system |
|---|---|---|---|---|
| Clinical Symptoms | ClinicalState | Psychomotor retardation | 1144814003 | Systematized Nomenclature of Medicine - Clinical Terms (SNOMED CT) |
| Clinical Symptoms | PersonCentered Disposition | Dyssomnia | 44186003 | Systematized Nomenclature of Medicine - Clinical Terms (SNOMED CT) |
| Clinical Symptoms | ClinicalState | Impaired concentration | 1144748009 | Systematized Nomenclature of Medicine - Clinical Terms (SNOMED CT) |
| Clinical Symptoms | PhysiologicalProcess | Weight loss | 816160009 | Systematized Nomenclature of Medicine - Clinical Terms (SNOMED CT) |
| Clinical Symptoms | PersonCentered Disposition | Suicidal thoughts | 6471006 | Systematized Nomenclature of Medicine - Clinical Terms (SNOMED CT) |
| Social Determinants | SocialRelationalQuality | Amount of workload | 10091a01 | Social Determinants of Health Ontology (SOHO) |

| | | | | |
|---|---|---|---|---|
| Social Determinants | SocialRelationalQuality | Perceived discrimination from society | 10063001 | Social Determinants of Health Ontology (SOHO) |
| Social Determinants | Psychological Disposition | Anxiety from community violence | 100710001 | Social Determinants of Health Ontology (SOHO) |
| Social Determinants | EconomicValueQuality | Lower earning | 100850001 | Social Determinants of Health Ontology (SOHO) |
| Social Determinants | Psychological Disposition | Financial stress | 10091601 | Social Determinants of Health Ontology (SOHO) |
| Trauma History | SocialCondition | Emotional Neglect | LP231633-1 | Adverse Childhood Experiences Ontology (ACESO) |
| Trauma History | SocialCondition | Suffered_From_Discrimination_Because_of_Race_Sexual_Orientation_Place_of_Birth_Disability_or_Religion | Suffered_From_Discrimination_Because_of_Race_Sexual_Orientation_Place_of_Birth_Disability_or_Religion | Adverse Childhood Experiences Ontology (ACESO) |
| Trauma History | SocialProcess | Lived with Parent or Guardian who Died | Lived_with_Parent_or_Guardian_who_Died | Adverse Childhood Experiences Ontology (ACESO) |
| Trauma History | SocialCondition | Mental Illness In Household | Mental_Illness_In_Household | Adverse Childhood Experiences Ontology (ACESO) |
| Trauma History | SocialProcess | Has Been Physically Abused | Has_Been_Physically_Abused | Adverse Childhood Experiences Ontology (ACESO) |
| Environmental Parameters | NaturalPhenotype Quality | Noise pollution | 8.7 | Sustainable cities and communities - Indicators for city services and quality of life (ISO 37120) |
| Environmental Parameters | NaturalPhenotype Quality | Fine particulate matter (PM2.5) concentration | 8.1 | Sustainable cities and communities - Indicators for city services and quality of life (ISO 37120) |
| Environmental Parameters | NaturalPhenotype Quality | Green area per 100,000 population | 19.1 | Sustainable cities and communities - Indicators for city services and quality of life (ISO 37120) |
| Environmental Parameters | TechnicalPerformance Quality | Annual number of public transport trips per capita | 18.3 | Sustainable cities and communities - Indicators for city services and quality of life (ISO 37120) |
| Environmental Parameters | NaturalPhenotype Quality | O3 (Ozone) concentration | 8.6 | Sustainable cities and communities - Indicators for city services and quality of life (ISO 37120) |
| Genetics and Molecular Biology | NaturalProcess | neurotransmitter transport | 0006836 | Gene Ontology (GO) |
| Genetics and Molecular Biology | NaturalProcess | catecholamine metabolic process | 0006584 | Gene Ontology (GO) |
| Genetics and Molecular Biology | NaturalProcess | negative regulation of neuron differentiation | 0045665 | Gene Ontology (GO) |
| Genetics and Molecular Biology | NaturalProcess | response to stress | 0006950 | Gene Ontology (GO) |
| Genetics and Molecular Biology | NaturalProcess | regulation of cytosolic calcium ion concentration | 0051480 | Gene Ontology (GO) |
| Physiology and Biomarkers | PhysiologicalQuality | Cortisol^AM peak specimen:MCnc:Pt:Ser/ | 9813-7 | Logical Observation Identifier Names and Codes (LOINC) |

| | | | | |
|---|---|---|---|---|
| | | Plas:Qn | | |
| Physiology and Biomarkers | PhysiologicalQuality | Heart rate:NRat:Pt:XXX:Qn | 8867-4 | Logical Observation Identifier Names and Codes (LOINC) |
| Physiology and Biomarkers | PhysiologicalRelational Quality | Body mass index:Ratio:Pt:^Patient:Qn | 39156-5 | Logical Observation Identifier Names and Codes (LOINC) |
| Physiology and Biomarkers | PhysiologicalQuality | Calcidiol+ercalcidiol:MCnc:Pt:Ser/Plas:Qn | 62292-8 | Logical Observation Identifier Names and Codes (LOINC) |
| Physiology and Biomarkers | PhysiologicalQuality | C reactive protein:MCnc:Pt:Ser/Plas:Qn | 1988-5 | Logical Observation Identifier Names and Codes (LOINC) |
| Behavioral and Lifestyle Data | Psychological Disposition | Detachment | P004R1 | Hierarchical Taxonomy of Psychopathology (HiTOP) |
| Behavioral and Lifestyle Data | Psychological Disposition | Disinhibition | P005xY | Hierarchical Taxonomy of Psychopathology (HiTOP) |
| Behavioral and Lifestyle Data | Psychological Disposition | Avolition | C00tUQ | Hierarchical Taxonomy of Psychopathology (HiTOP) |
| Behavioral and Lifestyle Data | Psychological Disposition | Anhedonia | C00XyI | Hierarchical Taxonomy of Psychopathology (HiTOP) |
| Behavioral and Lifestyle Data | Psychological Disposition | Antagonism | P006gM | Hierarchical Taxonomy of Psychopathology (HiTOP) |
| Neurocognitive and Psychometric Data | Psychological Disposition | Digit span reverse | C0589053 | Unified Medical Language System (UMLS) |
| Neurocognitive and Psychometric Data | Psychological Disposition | California verbal learning test | C0589055 | Unified Medical Language System (UMLS) |
| Neurocognitive and Psychometric Data | Psychological Disposition | Stroop Test | C2718024 | Unified Medical Language System (UMLS) |
| Neurocognitive and Psychometric Data | Psychological Disposition | TMT part A | C4063946 | Unified Medical Language System (UMLS) |
| Neurocognitive and Psychometric Data | Psychological Disposition | Wisconsin Card Sorting Test | C0451592 | Unified Medical Language System (UMLS) |
| Sensor Data | TechnicalProcess | Night-time screen activity | com.aware.provider.screen/screen | Smartphone sensing platform (AWARE) |
| Sensor Data | NaturalProcess | Circadian activity pattern | com.aware.provider.accelerometer.linear/sensor_linear_accelerometer | Smartphone sensing platform (AWARE) |
| Sensor Data | TechnicalProcess | Typing behavior | com.aware.provider.keyboard/keyboard | Smartphone sensing platform (AWARE) |
| Sensor Data | SocialProcess | Communication behavior | com.aware.provider.communication/calls | Smartphone sensing platform (AWARE) |
| Sensor Data | NaturalProcess | GPS-based mobility pattern | com.aware.provider.locations/locations | Smartphone sensing platform (AWARE) |
| Previous Diagnoses | Psychological Disposition | Social Phobias | F40.1 | International Classification of Diseases, Version 10 (ICD-10) |
| Previous Diagnoses | Psychological Disposition | Agoraphobia | F40.0 | International Classification of Diseases, Version 10 (ICD-10) |
| Previous Diagnoses | Psychological Disposition | Anankastic personality | F60.5 | International Classification of Diseases, |

| | | | | |
|---|---|---|---|---|
| | | disorder | | Version 10 (ICD-10) |
| Previous Diagnoses | ClinicalDisposition | Harmful Use of Alcohol | F10.1 | International Classification of Diseases, Version 10 (ICD-10) |
| Previous Diagnoses | Psychological Disposition | Somatization Disorder | F45.0 | International Classification of Diseases, Version 10 (ICD-10) |

**Table 4.** *Overview of the 50 heterogeneous data points integrated through the MMO. The table lists each data point together with its domain, corresponding MMO class, semantic code, and code system. This structured representation illustrates the traceability of mappings across medical, social, and behavioral domains and demonstrates the practical application of the MMO for standardized, interoperable patient data integration. Detailed justifications for the classification choices and their psychiatric relevance are provided in the supplementary materials.*

### 12.6 Semantic Classification and Psychiatric Relevance of Use Case Data Points

| Datapoint | Justification | Psychiatric relevance |
|---|---|---|
| **Clinical Symptoms:** Psychomotor retardation | The datapoint is classified in the Meta Mesh Ontology under ClinicalState because it denotes a continuant rather than an occurrent. In terms of the BFO/MMO hierarchy, it is not a transient process unfolding in time, but a relatively stable condition observable in a patient, situating it under continuant → sdc unit → ClinicalState. Its nature as a persisting manifestation distinguishes it from dispositions, which represent latent capacities or susceptibilities, and from processes, which capture dynamic events or changes. Psychomotor retardation instead reflects an ongoing clinical state, characterized by reduced motor and cognitive activity, that persists across time and provides the ontologically most accurate and consistent fit within ClinicalState. | It represents a marked slowing of thought, speech, and motor activity that directly reflects underlying disturbances in affective and neurocognitive processes. Clinically, its recognition is essential for the assessment and diagnosis of major depressive episodes, catatonia, and certain psychotic disorders, where it serves as a core symptom with diagnostic and prognostic weight. The presence and severity of psychomotor retardation provide interpretive value by indicating the extent of functional impairment, the depth of mood disturbance, and the likelihood of treatment resistance or suicidality, while its absence or reduction under therapy can signal clinical improvement. Thus, this datapoint is not merely descriptive but a critical clinical marker that integrates observable behavior with underlying psychopathology, shaping both diagnostic reasoning and therapeutic decision-making in psychiatry. |
| **Clinical Symptoms:** Dyssomnia | The datapoint is classified in the Meta Mesh Ontology under PersonCenteredDisposition because it refers to an enduring tendency of an individual to experience disturbances in sleep regulation rather than a transient state or unfolding process. Within the BFO/MMO hierarchy, this aligns with continuant → sdc unit → realizable unit → disposition unit → PersonCenteredDisposition, since it captures a realizable entity that inheres in a person and may | It encompasses disturbances in the quantity, quality, or timing of sleep that are frequently intertwined with mental disorders. Clinically, the identification of dyssomnia is crucial because disordered sleep often serves both as a symptom and as a risk factor in conditions such as major depressive disorder, generalized anxiety disorder, bipolar disorder, and post-traumatic stress disorder, influencing both diagnostic classification and treatment strategies. Its presence, persistence, or severity provides interpretive value by signaling underlying affective dysregulation, cognitive |

| | | |
|---|---|---|
| | be manifested under certain conditions. Its defining features show that it is not simply a momentary clinical state, which would imply an observable condition at a given time, nor a process, which would involve the dynamic unfolding of sleep-related behaviors. Instead, Dyssomnia expresses a persistent susceptibility of the person to altered sleep patterns, which may manifest variably but reflects a deeper dispositional structure. This ontological positioning provides the most accurate abstraction level, situating Dyssomnia as a person-centered disposition that explains and grounds the recurrent clinical observations associated with sleep dysfunction. | impairments, or heightened stress reactivity, while its absence or resolution under therapy may indicate stabilization of the broader psychopathological state. Consequently, dyssomnia functions as a sensitive clinical marker that bridges subjective patient experience with objective psychopathological processes, guiding comprehensive psychiatric assessment and informing tailored therapeutic interventions. |
| **Clinical Symptoms:** Impaired concentration | The datapoint is classified in the Meta Mesh Ontology under ClinicalState because it denotes a persisting condition observable in a patient rather than a transient event or a latent capacity. Within the BFO/MMO hierarchy, this aligns with continuant → sdc unit → ClinicalState, as it characterizes a relatively stable alteration in cognitive function that can be clinically assessed at a given time. Its definitional features show that it does not describe a process, since it lacks the dynamic unfolding characteristic of occurrents, nor does it represent a disposition, as it does not capture a latent tendency or susceptibility but rather a manifest condition of diminished attentional performance. By being a temporally extended yet immediately observable state, impaired concentration is ontologically most consistent with the class of ClinicalState. | It reflects disturbances in attentional control and cognitive processing that are integral to the assessment of numerous psychiatric conditions. Clinically, its identification is particularly significant in the evaluation of depressive disorders, anxiety disorders, attention-deficit/hyperactivity disorder, and psychotic spectrum conditions, where concentration deficits often serve as both diagnostic criteria and therapeutic targets. The presence and degree of impaired concentration provide interpretive value by indicating the extent of functional impairment, the severity of mood or cognitive dysregulation, and the impact of psychopathology on daily living, while its improvement or resolution may serve as a reliable indicator of treatment response. As such, this symptom operates as a crucial bridge between subjective patient experience and measurable cognitive dysfunction, thereby supporting accurate diagnosis, monitoring of disease trajectory, and the tailoring of individualized treatment in psychiatric practice. |
| **Clinical Symptoms:** Weight loss | The datapoint is classified in the Meta Mesh Ontology under PhysiologicalProcess because it denotes a dynamic and temporally extended occurrence rather than a static state or latent disposition. Within the BFO/MMO hierarchy, this aligns with occurent unit → process unit → HealthProcess → PhysiologicalProcess, since weight loss unfolds through continuous metabolic and bodily changes that reduce body mass over time. Its characteristics show that it is not adequately represented as a clinical state, which would capture an immediately observable condition at a given point in time, nor as a disposition, which would imply a latent tendency that may or may not manifest. Instead, weight loss exemplifies a physiological process in which | It frequently reflects underlying disturbances in mood, cognition, or behavior that are characteristic of a range of psychiatric disorders. Clinically, unintentional weight loss is a critical symptom in major depressive disorder, anorexia nervosa, and other eating disorders, as well as in certain psychotic and anxiety conditions, where it may signify maladaptive alterations in appetite regulation, motivation, or interoceptive awareness. Its presence and severity provide interpretive value by indicating the degree of physiological compromise, the intensity of psychopathological processes, and the potential risk of medical complications, while its absence or reversal may mark therapeutic progress or stabilization of the psychiatric condition. Thus, weight loss functions not only as a somatic manifestation of mental illness but also as a measurable indicator of psychiatric burden and treatment response, reinforcing its role as an |

| | | |
|---|---|---|
| | energy balance, nutrition, and metabolism interact dynamically to produce a measurable outcome, making PhysiologicalProcess the most accurate ontological classification. | essential parameter in comprehensive psychiatric assessment and care. |
| **Clinical Symptoms:** Suicidal thoughts | The datapoint is classified in the Meta Mesh Ontology under PersonCenteredDisposition because it captures a realizable tendency that inheres in a person and may manifest in the form of recurring ideation or intent under specific psychological or contextual conditions. Within the BFO/MMO hierarchy, this corresponds to continuant → sdc unit → realizable unit → disposition unit → PersonCenteredDisposition, since it is not an immediately observable static state but rather a persistent capacity-like orientation of the individual toward self-destructive thinking. Its characteristics show that it cannot be adequately modeled as a clinical state, which would imply a directly present and temporally bounded condition, nor as a process, which would involve the unfolding of actions or events in time. Instead, suicidal thoughts exemplify a dispositional structure grounded in the person, capable of being actualized in episodes of ideation but persisting even when not actively expressed, thereby justifying its placement as a PersonCenteredDisposition. | They directly reflect disturbances in mood, cognition, and existential appraisal that place the individual at heightened risk of self-harm or suicide. Clinically, their recognition is essential in the assessment of major depressive disorder, bipolar disorder, psychotic disorders, and certain personality disorders, where suicidal ideation serves as a critical indicator of severity and an urgent determinant of treatment planning. The presence, persistence, or intensity of suicidal thoughts provides interpretive value by signaling the degree of psychological distress, hopelessness, and impaired coping, while their absence or reduction under therapy may reflect meaningful stabilization or recovery. As such, suicidal thoughts function not merely as a symptom but as an immediate marker of psychiatric risk that necessitates comprehensive evaluation, close monitoring, and targeted intervention, underscoring their central role in both diagnosis and clinical management within psychiatry. |
| **Social Determinants:** Amount of workload | The datapoint is classified in the Meta Mesh Ontology under SocialRelationalQuality because it expresses a quality that inheres in the structured relations between a person and their social or occupational environment rather than in the individual alone. Within the BFO/MMO hierarchy, this corresponds to continuant → sdc unit → quality unit → relational quality unit → SocialRelationalQuality, since workload is not an isolated property but a relational measure of demands placed upon an individual within a social system. Its characteristics show that it is not a disposition, which would imply a latent capacity or susceptibility within the person, nor a process, which would describe the dynamic unfolding of tasks over time. Instead, the amount of workload captures a relational quality defined through the interaction between individual and social structures, making its placement under SocialRelationalQuality ontologically coherent and precise. | The quantity and intensity of occupational or academic demands exert a profound influence on mental health and resilience. Clinically, workload assessment is essential in understanding the role of chronic stress exposure, overcommitment, or insufficient recovery time in the onset and maintenance of conditions such as burnout, major depressive disorder, anxiety disorders, and stress-related somatic syndromes. The presence of excessive workload can serve as a marker of heightened psychosocial strain, maladaptive coping, and increased vulnerability to psychiatric morbidity, while a markedly reduced or absent workload may reveal social withdrawal, impaired functioning, or the psychiatric sequelae of illness that limit occupational participation. The severity and trajectory of workload therefore provide interpretive value for evaluating both risk factors and functional outcomes, supporting comprehensive psychiatric assessment and guiding targeted interventions that address not only symptoms but also the broader social context of mental health. |

| | | |
|---|---|---|
| **Social Determinants:** Perceived discrimination from society | The datapoint is classified in the Meta Mesh Ontology under SocialRelationalQuality because it denotes a quality that arises within the relational dynamics between an individual and the broader social structures in which they are embedded. Within the BFO/MMO hierarchy, this aligns with continuant → sdc unit → quality unit → relational quality unit → SocialRelationalQuality, since discrimination is not an intrinsic feature of the individual but a socially mediated quality that reflects how the person is positioned and treated within collective interactions. Its nature excludes classification as a disposition, because it does not express a latent capacity or inherent tendency of the person, and it is also not a process, as it does not primarily describe a dynamic sequence of events but rather the relational quality that persists and characterizes the individual's lived experience of societal bias. By capturing this structured relation as a quality inhering in the social bond between person and society, the datapoint is most coherently and precisely situated under SocialRelationalQuality. | It represents a persistent psychosocial stressor that shapes individual vulnerability to mental illness and influences the course of psychiatric disorders. Clinically, the assessment of perceived discrimination is critical for understanding how experiences of social exclusion, stigma, or structural inequity contribute to the onset and maintenance of depressive, anxiety, psychotic, and trauma-related disorders, as well as to maladaptive coping strategies such as substance use. The presence and severity of perceived discrimination provide interpretive value by indicating cumulative stress exposure, diminished social support, and potential barriers to treatment engagement, while its absence or mitigation may reflect protective psychosocial resources that buffer psychiatric risk. Thus, this datapoint functions as a key indicator linking social context to individual psychopathology, enabling more accurate psychiatric assessment and guiding interventions that integrate clinical care with strategies to address social adversity. |
| **Social Determinants:** Anxiety from community violence | The datapoint is classified in the Meta Mesh Ontology under PsychologicalDisposition because it denotes a realizable tendency of an individual's psychological makeup that may be actualized in response to environmental triggers such as exposure to violence in the community. Within the BFO/MMO hierarchy, this aligns with continuant → sdc unit → realizable unit → disposition unit → PersonCenteredDisposition → PsychologicalDisposition, since it inheres in the person as a persisting susceptibility to manifest anxious responses rather than as a directly observable state or unfolding event. Its characteristics show that it is not a clinical state, which would imply a currently present condition of anxiety at a given moment, nor a process, which would capture the dynamic sequence of fear reactions over time. Instead, it describes an enduring psychological disposition that grounds the potential for anxiety episodes when the relevant social context is present, making PsychologicalDisposition the most ontologically accurate classification. | Exposure to violence in one's social environment constitutes a potent stressor that heightens vulnerability to a wide range of mental health disorders. Clinically, assessing anxiety related to community violence is essential for identifying individuals at risk of post-traumatic stress disorder, generalized anxiety disorder, depressive syndromes, and substance use disorders, as well as for understanding the cumulative impact of chronic fear and hypervigilance on psychological functioning. The presence and intensity of such anxiety provide interpretive value by indicating the degree of trauma-related symptomatology, impaired sense of safety, and diminished social trust, while its absence or reduction may suggest resilience, effective coping mechanisms, or recovery following targeted intervention. However, changes in anxiety levels may also reflect alterations in the external environment - such as relocation, reduced exposure to violence, or temporary protective contexts - highlighting the complex interaction between individual and contextual determinants of psychological adaptation. As such, this datapoint serves as a critical marker linking environmental adversity to psychiatric morbidity, supporting more comprehensive assessment and guiding treatment strategies that address both individual symptoms and the broader social context of violence exposure. |

| | | |
|---|---|---|
| **Social Determinants:** Lower earning | The datapoint is classified in the Meta Mesh Ontology under EconomicValueQuality because it designates a quality that inheres in the economic situation of an individual or household and expresses the relative valuation of income within a social and economic system. Within the BFO/MMO hierarchy, this corresponds to continuant → sdc unit → quality unit → EconomicValueQuality, since earnings are not intrinsic objects or processes but evaluative measures of worth situated in economic relations. Its nature excludes interpretation as a disposition, because it does not describe a latent potential or capacity of a person, and it is not a process, as it does not denote the unfolding activity of generating income but rather the comparative level of value attached to those earnings at a given time. By capturing the enduring evaluative aspect of economic standing, "Lower earning" is most coherently placed under EconomicValueQuality, ensuring an ontologically precise representation. | Economic disadvantage exerts a profound influence on both the development and course of mental disorders. Clinically, the assessment of reduced income is essential for understanding barriers to accessing care, medication adherence, and psychosocial stability, as financial strain is strongly associated with elevated rates of depression, anxiety, substance use, and stress-related disorders. The presence and severity of lower earning provide interpretive value by reflecting chronic stress exposure, reduced autonomy, and diminished coping resources, while its absence or improvement may signal enhanced resilience, social mobility, or recovery processes that support mental health stabilization. Thus, this datapoint functions not only as an indicator of socioeconomic risk but also as a dynamic marker of psychiatric vulnerability and recovery potential, underscoring the importance of integrating social determinants into comprehensive psychiatric assessment and treatment planning. |
| **Social Determinants:** Financial stress | The datapoint is classified in the Meta Mesh Ontology under PsychologicalDisposition because it denotes an enduring psychological tendency to experience strain, worry, or anxiety in response to financial hardship. Within the BFO/MMO hierarchy, this corresponds to continuant → sdc unit → realizable unit → disposition unit → PersonCenteredDisposition → PsychologicalDisposition, as the phenomenon inheres in the individual as a latent vulnerability that becomes manifest under certain socio-economic conditions. It is not appropriately modeled as an economic quality, since the defining feature of the datapoint is not the objective financial status itself but the psychological orientation towards it. Nor is it a process, which would imply the unfolding temporal dynamics of a stress episode, while "financial stress" designates a persistent dispositional structure that grounds the possibility of such episodes. By situating the phenomenon as a realizable psychological disposition that links external economic challenges to internal mental responses, its classification under PsychologicalDisposition ensures ontological precision and alignment with the BFO/MMO framework. | Persistent economic strain constitutes a well-established risk factor for the onset, exacerbation, and chronicity of mental disorders. Clinically, the evaluation of financial stress is crucial for understanding how economic insecurity, debt burden, or instability in meeting basic needs contributes to depressive disorders, generalized anxiety, substance misuse, and suicidality, as well as for identifying potential barriers to treatment adherence and recovery. The presence and intensity of financial stress provide interpretive value by signaling heightened psychosocial vulnerability, impaired coping capacity, and an elevated risk of functional decline, whereas its absence or resolution may reflect protective resources, improved social support, or recovery trajectories that strengthen psychiatric resilience. Thus, this datapoint serves as a critical link between socioeconomic adversity and individual psychopathology, guiding comprehensive psychiatric assessment and informing integrative interventions that address both clinical symptoms and underlying social stressors. |
| **Trauma History:** | The datapoint is classified in the Meta Mesh | It reflects the absence of adequate emotional support, |

| | | |
|---|---|---|
| Emotional Neglect | Ontology under SocialCondition because it denotes a persisting socio-relational circumstance characterized by the absence of expected emotional care within a given interpersonal or familial context. Within the BFO/MMO hierarchy, this corresponds to continuant → sdc unit → SocialCondition, since emotional neglect is not an event unfolding in time but a condition that endures as a stable aspect of the social environment in which an individual develops. It is not adequately captured as a disposition, because it does not describe a latent capacity within an individual, but rather an externally situated state that constrains or shapes relational experiences. Likewise, it is not a process, which would denote discrete temporal episodes of neglectful interactions, whereas the construct refers to the condition of being subjected to such neglect as an enduring social reality. Its classification as a SocialCondition therefore provides ontological precision by situating the phenomenon as a structural, context-dependent factor that shapes an individual's social and psychological development within the MMO framework. | validation, and responsiveness by caregivers during a developmental period that significantly shapes long-term mental health outcomes. Clinically, the assessment of emotional neglect is crucial for understanding vulnerability to depressive disorders, anxiety disorders, personality pathology, post-traumatic stress disorder, and difficulties in attachment and emotional regulation, as such early adverse experiences frequently disrupt core processes of self-concept and interpersonal functioning. The presence and severity of emotional neglect provide interpretive value by indicating the depth of developmental trauma, the likelihood of maladaptive coping strategies, and the persistence of relational difficulties that contribute to ongoing psychiatric morbidity, while its absence or mitigation through protective caregiving may suggest resilience and a reduced risk for psychopathology. Thus, this datapoint functions as a critical marker of early psychosocial adversity that informs both diagnostic formulation and therapeutic intervention, highlighting the enduring influence of trauma history on psychiatric assessment and treatment planning. |
| **Trauma History:** Suffered from discrimination because of race, sexual orientation, place of birth, disability, or religion | The datapoint is classified in the Meta Mesh Ontology under SocialCondition because it refers to a persisting socially mediated circumstance in which individuals are subjected to structurally embedded disadvantage and exclusion based on identity characteristics. Within the BFO/MMO hierarchy, this corresponds to continuant → sdc unit → SocialCondition, since discrimination in this sense is not reducible to a transient event or isolated interaction but constitutes an enduring condition of the social environment that shapes the lived experiences of the affected person. It cannot be modeled as a disposition, because it does not denote an internal capacity or latent tendency inhering in the individual, and it is not a process, since it does not primarily describe the temporal unfolding of discriminatory acts but rather the stable social condition of being subjected to systematic bias. Framing it as a SocialCondition thus provides ontological accuracy by situating discrimination as a structural and relational circumstance that persists within the social domain, capturing its contextual and enduring nature within the MMO framework. | It represents a form of chronic psychosocial trauma with lasting impact on identity, self-worth, and mental health. Clinically, the evaluation of such discrimination is essential for understanding pathways to depressive disorders, anxiety, post-traumatic stress disorder, and substance use disorders, as well as the development of internalized stigma and mistrust that may interfere with therapeutic alliance and treatment adherence. The presence and severity of this psychosocial stressor provide interpretive value by signaling cumulative stress exposure, heightened vulnerability to psychopathology, and the risk of social isolation or maladaptive coping strategies, whereas its absence or mitigation through supportive environments may indicate protective social factors that foster resilience. Thus, this datapoint functions as a crucial indicator of trauma rooted in systemic inequities, linking broader sociocultural adversity to individual psychiatric outcomes and guiding both diagnostic formulation and culturally sensitive interventions. |

| | | |
|---|---|---|
| **Trauma History:** Lived with parent or guardian who died | The datapoint is classified in the Meta Mesh Ontology under SocialProcess because it denotes a temporally extended event situated within a social context, namely the familial relationship between a child and a caregiver. Within the BFO/MMO hierarchy, this corresponds to entity → occurrent unit → process unit → SocialProcess, as it unfolds over time and consists of the experiential trajectory of cohabitation culminating in the death of the parent or guardian. This classification distinguishes the phenomenon from continuant categories, since it cannot be adequately represented as a persisting condition or disposition that inheres in the individual, but rather as an eventive occurrence that is socially mediated and relationally structured. It is not a PsychologicalDisposition, since it does not describe an enduring internal tendency or latent capacity, but the lived social circumstance of loss within a familial household. Nor is it a SocialCondition, because it does not designate a stable contextual circumstance but an unfolding transition in the social fabric of the individual's life. By modeling it as a SocialProcess, the MMO captures its ontological status as a temporally structured, socially embedded event that plays a formative role in shaping trauma histories. | Early or close exposure to the death of a primary caregiver constitutes a profound disruption in attachment, stability, and emotional security, all of which are foundational to psychological development. Clinically, the assessment of this experience is critical for identifying vulnerability to depressive and anxiety disorders, post-traumatic stress disorder, complicated grief, and maladaptive attachment patterns that may persist into adulthood. The presence and severity of this traumatic event provide interpretive value by indicating heightened risk for difficulties in emotional regulation, trust, and interpersonal functioning, while its absence or the presence of mitigating protective factors, such as supportive caregiving after the loss, may signal greater resilience and reduced psychiatric vulnerability. Thus, this datapoint serves as an essential marker of developmental trauma that informs diagnostic understanding, contextualizes symptomatology, and guides treatment strategies that integrate grief processing and relational repair into psychiatric care. |
| **Trauma History:** Mental illness in household | The datapoint is classified in the Meta Mesh Ontology under SocialCondition because it denotes a persisting contextual circumstance that characterizes the social environment of the individual rather than a transient event or an inherent quality of the person. Within the BFO/MMO hierarchy, this aligns with entity → continuant unit → sdc unit → SocialCondition, since the phenomenon reflects a relatively stable social reality in which the presence of mental illness within a household structures the lived conditions of its members. It cannot be modeled as a disposition, because it does not describe a capacity or tendency inhering in an individual but rather a relational and environmental context. Nor does it qualify as a process, since it does not primarily denote a temporally unfolding sequence of events but instead a standing condition of the social milieu. By situating it as a SocialCondition, the MMO framework captures its ontological status as a continuing circumstance that exerts | Growing up or living in an environment affected by psychiatric morbidity profoundly shapes emotional development, coping mechanisms, and vulnerability to mental disorders. Clinically, documenting such a history is essential for understanding intergenerational transmission of psychiatric risk, exposure to dysfunctional family dynamics, and the potential normalization of maladaptive behaviors that may contribute to later psychopathology, including depressive disorders, anxiety, substance use, and relational difficulties. The presence and severity of this factor provide interpretive value by highlighting cumulative stress exposure, impaired caregiving consistency, and potential modeling of maladaptive coping strategies, while its absence may indicate a more stable psychosocial environment that fosters resilience. Thus, this datapoint functions as a critical contextual marker linking familial psychiatric burden to individual mental health trajectories, thereby informing diagnostic formulation, risk assessment, and the design of interventions that integrate both clinical care and family support. |

| | | |
|---|---|---|
| | formative influence on developmental and psychosocial outcomes while maintaining its distinction from both intrapersonal dispositions and eventive occurrences. | |
| **Trauma History:** Has been physically abused | The datapoint is classified in the Meta Mesh Ontology under SocialProcess because it designates an eventive phenomenon that unfolds through interpersonal interaction and involves the infliction of harm within a social context. Within the BFO/MMO hierarchy, this corresponds to entity → occurrent unit → process unit → SocialProcess, since the experience of physical abuse is not a static condition or internal capacity but a temporally extended process that occurs between agents in a structured social environment. It cannot be adequately represented as a disposition, because it does not describe a latent tendency inhering in an individual, nor as a SocialCondition, since it is not a persisting contextual circumstance but rather a concrete series of harmful acts. Modeling it as a SocialProcess ensures ontological precision by capturing its status as a relationally mediated, temporally unfolding event that contributes to the shaping of trauma histories. | Exposure to physical violence constitutes a severe adverse experience with enduring effects on psychological functioning, emotional regulation, and interpersonal trust. Clinically, identifying a history of physical abuse is essential for assessing risk of post-traumatic stress disorder, depressive and anxiety disorders, substance use, and personality pathology, as well as for understanding patterns of hypervigilance, aggression, or dissociation that may complicate diagnosis and treatment. The presence and severity of physical abuse provide interpretive value by indicating the depth of traumatic imprint, the likelihood of maladaptive coping strategies, and heightened vulnerability to revictimization or self-destructive behavior, while its absence or mitigation through protective factors such as stable support networks may suggest greater resilience and adaptive capacity. Thus, this datapoint functions as a key marker of trauma-related psychopathology, guiding comprehensive psychiatric assessment and informing therapeutic interventions that must integrate trauma-focused care to address both symptomatology and underlying developmental wounds. |
| **Environmental Parameters:**<br><br>Noise pollution | The datapoint is classified in the Meta Mesh Ontology under NaturalPhenotypeQuality because it expresses a measurable quality that inheres in the natural environment and is manifested as a persistent characteristic of a physical setting. Within the BFO/MMO hierarchy, this corresponds to entity → continuant unit → sdc unit → quality unit → NaturalPhenotypeQuality, since noise pollution is neither an independent material entity nor an immaterial site, but a quality borne by environmental regions that reflects the intensity and distribution of sound as a natural phenomenon. It cannot be represented as a disposition, because it does not describe a latent capacity or potential of an entity, nor as a process, since it does not denote a temporally unfolding sequence of events but rather a persisting qualitative state of the environment at any given time. Classifying it as a NaturalPhenotypeQuality provides ontological precision by situating it as an objectively measurable quality of environmental systems, aligning its abstraction level with other naturally occurring phenotype-level conditions that describe | Chronic exposure to excessive environmental noise constitutes a persistent stressor that adversely affects cognitive functioning, sleep regulation, and emotional stability. Clinically, the evaluation of noise pollution is important for understanding its role in the exacerbation of psychiatric conditions such as generalized anxiety disorder, major depressive disorder, insomnia, and stress-related syndromes, as well as for recognizing its contribution to irritability, impaired concentration, and diminished quality of life. The presence and severity of noise pollution provide interpretive value by signaling heightened physiological arousal, sleep disruption, and cumulative stress burden, while its absence or reduction may reflect protective environmental conditions that support psychological resilience and recovery. Thus, this datapoint serves as a critical link between environmental adversity and psychiatric morbidity, highlighting the necessity of integrating ecological factors into comprehensive psychiatric assessment and treatment planning. |

| | | |
|---|---|---|
| | the state of ecological or urban habitats. | |
| **Environmental Parameters:**<br><br>Fine particulate matter (PM2.5) concentration | The datapoint is classified in the Meta Mesh Ontology under NaturalPhenotypeQuality because it denotes a measurable, inhering quality of the natural environment that expresses the concentration of particulate matter in ambient air. Within the BFO/MMO hierarchy, this corresponds to entity → continuant unit → sdc unit → quality unit → NaturalPhenotypeQuality, since PM2.5 concentration is not an independent object or site but a quality borne by atmospheric regions that can be continuously measured and compared. It cannot be represented as a disposition, because it does not describe a latent capacity or potentiality of an entity, but rather a realized, observable state of environmental air. Nor does it correspond to a process, since it does not primarily capture the dynamic unfolding of particulate emissions or dispersion over time, but rather the persisting condition of air quality at a given point. By classifying it as a NaturalPhenotypeQuality, the MMO framework ensures ontological precision by situating it as an intrinsic, phenotype-level quality of natural systems, aligning with the measurement-based representation of environmental states required for sustainability and public health assessments. | Chronic exposure to elevated air pollution has been consistently associated with neuroinflammation, oxidative stress, and vascular dysfunction, all of which contribute to the onset and progression of psychiatric disorders. Clinically, assessing PM2.5 levels is important for understanding environmental contributions to depression, anxiety, cognitive decline, and neurodevelopmental conditions, as well as for identifying modifiable risk factors that exacerbate vulnerability in already affected individuals. The presence and severity of elevated PM2.5 concentrations provide interpretive value by indicating an increased burden of environmental stress that may manifest as mood disturbances, cognitive impairment, or heightened susceptibility to psychiatric relapse, while low or absent exposure may reflect protective conditions that support mental health stability and recovery. Thus, this datapoint functions as a vital ecological indicator linking environmental quality to psychiatric morbidity, reinforcing the need to integrate environmental determinants into comprehensive psychiatric assessment and intervention strategies. |
| **Environmental Parameters:**<br><br>Green area per 100,000 population | The datapoint is classified in the Meta Mesh Ontology under NaturalPhenotypeQuality because it denotes a measurable quality of the natural environment that persists as a structural feature of the urban or regional landscape. Within the BFO/MMO hierarchy, this corresponds to entity → continuant unit → sdc unit → quality unit → NaturalPhenotypeQuality, since the extent of accessible green space is not itself an independent object or aggregate, but a quantifiable property inhering in natural sites and regions. It is not appropriately represented as a disposition, because it does not describe a latent potential or capacity of the environment, but instead reflects an actualized, observable state of ecological provision within a given population context. Likewise, it cannot be categorized as a process, since it does not capture the unfolding of land-use change or vegetation growth, but rather the persisting condition of spatial availability of natural areas relative to human population density. Classifying it as a | Access to natural spaces has been shown to buffer stress, enhance emotional regulation, and promote cognitive restoration. Clinically, evaluating the availability of green areas is important for understanding environmental contributions to the prevalence and severity of depressive and anxiety disorders, stress-related conditions, and cognitive fatigue, as well as for identifying protective factors that support recovery and resilience. The presence and adequacy of green space provide interpretive value by indicating opportunities for physical activity, social interaction, and restorative experiences that mitigate psychiatric symptomatology, while its absence or scarcity may reflect increased risk of chronic stress, social isolation, and heightened vulnerability to mental illness. Thus, this datapoint serves as an ecological marker that directly links urban design and environmental quality to psychiatric well-being, underscoring the importance of integrating environmental determinants into comprehensive psychiatric assessment and public health planning. |

| | | |
|---|---|---|
| | NaturalPhenotypeQuality ensures ontological precision by situating it as an intrinsic, phenotype-level environmental quality, which provides a stable basis for assessing urban sustainability and public health indicators. | |
| **Environmental Parameters:**<br><br>Annual number of public transport trips per capita | The datapoint is classified in the Meta Mesh Ontology under TechnicalPerformanceQuality because it expresses a measurable quality of the functioning of a technical system, namely the public transport infrastructure, in relation to population use. Within the BFO/MMO hierarchy, this aligns with entity → continuant unit → sdc unit → quality unit → TechnicalPerformanceQuality, since the indicator denotes an evaluative property that inheres in the operational performance of a transport system rather than in independent objects or social aggregates. It is not appropriately modeled as a disposition, as it does not represent a latent capacity of the system to transport passengers, but rather an empirically realized and continuously measurable output of system performance over time. Nor does it fall under the category of process, because the indicator does not describe the unfolding of individual journeys or transport events, but instead captures the enduring, quantifiable quality of system utilization relative to population size. By situating this datapoint as a TechnicalPerformanceQuality, the ontology ensures conceptual precision by recognizing it as an intrinsic quality of a technical system that serves as a benchmark for efficiency, accessibility, and sustainability assessments in urban environments. | Patterns of mobility and accessibility strongly influence social participation, occupational functioning, and exposure to environmental stressors. Clinically, evaluating public transport use is important for understanding indirect determinants of mental health, as reliable and safe transport facilitates access to healthcare, social networks, and employment, while limited or stressful commuting conditions have been linked to increased rates of anxiety, depression, and perceived stress. The presence of frequent and accessible transport use may indicate greater social integration, autonomy, and protective environmental conditions, whereas low usage or constrained access can reveal social isolation, economic barriers, or functional impairments associated with psychiatric morbidity. Thus, this datapoint functions as a valuable ecological indicator that connects patterns of urban infrastructure and mobility with psychiatric well-being, supporting integrative assessment and informing interventions that address both clinical needs and broader determinants of mental health. |
| **Environmental Parameters:**<br><br>O3 (Ozone) concentration | The datapoint is classified in the Meta Mesh Ontology under NaturalPhenotypeQuality because it denotes a quantifiable, intrinsic quality of the natural atmosphere that persists as a measurable environmental condition. Within the BFO/MMO hierarchy, this corresponds to entity → continuant unit → sdc unit → quality unit → NaturalPhenotypeQuality, since ozone concentration is not an independent material object but rather a quality inhering in a natural region of air, which can be empirically assessed through environmental monitoring. It cannot be modeled as a disposition, because it does not describe a latent potential or capacity of the atmosphere but instead | Elevated levels of ambient ozone have been linked to neuroinflammatory processes, oxidative stress, and impaired neurocognitive functioning, all of which contribute to psychiatric vulnerability. Clinically, assessing ozone exposure is important for understanding environmental contributions to the incidence and severity of depression, anxiety, cognitive decline, and neurodevelopmental disorders, particularly in urban populations with sustained exposure. The presence and severity of elevated $O_3$ concentrations provide interpretive value by indicating an increased environmental burden that may exacerbate mood disturbances, impair attention and memory, and elevate stress sensitivity, while lower or controlled levels may reflect protective ecological conditions that support |

| | | |
|---|---|---|
| | reflects an actually instantiated state of chemical composition in a given place and time. Nor does it correspond to a process, as the indicator does not capture the dynamic photochemical reactions that generate ozone, but rather the persisting measurable condition of air quality at a specific concentration level. Classifying it as a NaturalPhenotypeQuality thus provides ontological precision by situating it as an inherent, phenotype-level environmental quality of atmospheric systems, consistent with the need for stable and comparable indicators in sustainability and public health frameworks. | psychological stability and recovery. Thus, this datapoint serves as a critical environmental marker that connects air quality to psychiatric outcomes, emphasizing the necessity of integrating ecological risk factors into comprehensive psychiatric assessment and preventive strategies. |
| **Genetics and Molecular Biology:**<br><br>neurotransmitter transport | The datapoint is classified in the Meta Mesh Ontology under NaturalProcess because it denotes a temporally extended, dynamic event in which neurotransmitter molecules are actively moved across cellular compartments or synaptic boundaries. Within the BFO/MMO hierarchy, this corresponds to entity → occurent unit → process unit → NaturalProcess, since the transport activity is not a static continuant but an unfolding biological phenomenon that depends on causal interactions between cellular structures and molecular agents. It cannot be represented as a disposition, because it does not signify a latent capacity of a neuron or membrane protein, but rather the actualized occurrence of translocation in space and time. Nor is it appropriately modeled as a quality, since it is not a persisting state or measurable property of an entity but an eventive activity characterized by change and directedness. By situating this datapoint as a NaturalProcess, the ontology maintains ontological coherence by recognizing neurotransmitter transport as a fundamental physiological process intrinsic to neural systems and essential to signaling dynamics. | It governs the regulation, release, and reuptake of key neurochemicals that underlie synaptic transmission and neural circuit functioning. Clinically, the integrity of neurotransmitter transport systems is critical for understanding the pathophysiology of mood disorders, psychotic disorders, anxiety disorders, and neurodevelopmental conditions, as dysregulation in serotonin, dopamine, or glutamate transport has been directly implicated in symptom expression and treatment response. The presence of aberrant neurotransmitter transport activity provides interpretive value by signaling altered synaptic communication, impaired neuroplasticity, and disrupted affective or cognitive processing, while normalized or therapeutically modulated transport can serve as a biomarker of clinical improvement. Thus, this datapoint functions as a molecular-level determinant of psychiatric morbidity and recovery, bridging genetic and neurobiological mechanisms with observable clinical phenomena and guiding both pharmacological and precision medicine approaches in psychiatry. |
| **Genetics and Molecular Biology:**<br><br>catecholamine metabolic process | The datapoint is classified in the Meta Mesh Ontology under NaturalProcess because it refers to a temporally extended sequence of biochemical transformations by which catecholamines are synthesized, modified, or degraded within living systems. Within the BFO/MMO hierarchy, this is situated as entity → occurent unit → process unit → NaturalProcess, since the metabolic pathway is not a static continuant but a dynamic occurrence that unfolds through enzymatic activity in space | It encompasses the synthesis, degradation, and regulation of neurotransmitters such as dopamine, norepinephrine, and epinephrine, which are central to mood, cognition, and stress response. Clinically, disturbances in catecholamine metabolism are implicated in the pathophysiology of major depressive disorder, bipolar disorder, schizophrenia, attention-deficit/hyperactivity disorder, and anxiety disorders, making it a key focus for both diagnostic consideration and therapeutic targeting. The presence of dysregulated catecholamine metabolism provides |

| | | |
|---|---|---|
| | and time. It cannot be appropriately represented as a disposition, because it does not signify a latent capacity of a cell or enzyme but instead designates the realized series of metabolic events. Nor is it accurately modeled as a quality, since the focus is not on a persisting state or measurable property but on the unfolding biochemical process itself. Classifying it as a NaturalProcess thus captures its ontological status as a biological phenomenon that is both intrinsic to natural systems and temporally structured, aligning with the MMO's systematic treatment of molecular and physiological processes as occurents. | interpretive value by indicating altered neurotransmitter availability, impaired stress reactivity, and vulnerability to affective or cognitive dysfunction, while normalization through pharmacological or biological interventions often corresponds with symptomatic improvement. Thus, this datapoint functions as a critical molecular marker linking neurochemical regulation to psychiatric states, guiding precision assessment and informing treatment strategies that address the biochemical underpinnings of mental illness. |
| **Genetics and Molecular Biology:**<br><br>negative regulation of neuron differentiation | The datapoint is classified in the Meta Mesh Ontology under NaturalProcess because it denotes a temporally extended biological phenomenon in which molecular and cellular mechanisms act to decrease or inhibit the progression of precursor cells into differentiated neurons. Within the BFO/MMO hierarchy, this aligns with entity → occurent unit → process unit → NaturalProcess, since the regulation described is not a static continuant but an eventive unfolding embedded in developmental biology. It cannot be categorized as a disposition, because it does not describe a latent potential or capacity residing in a cell or gene product, but rather the realized enactment of inhibitory mechanisms over time. Nor is it adequately modeled as a quality, since it is not a persisting property inhering in an entity but instead a directed, causal sequence of molecular interactions with temporal extension. Classifying this datapoint as a NaturalProcess preserves ontological coherence by recognizing it as a concrete biological occurrence situated in the regulation of developmental pathways, thereby distinguishing it from static attributes or unrealized capacities. | It reflects molecular and cellular mechanisms that control the maturation and specialization of neurons, processes fundamental to brain development and plasticity. Clinically, disturbances in this regulatory pathway are associated with neurodevelopmental and psychiatric disorders such as autism spectrum disorder, schizophrenia, and major depressive disorder, where impaired neuronal differentiation can disrupt cortical connectivity, synaptic integration, and functional brain circuitry. The presence of abnormal or excessive negative regulation provides interpretive value by indicating potential deficits in neurogenesis and adaptive plasticity, contributing to cognitive dysfunction, affective instability, and altered stress reactivity, while balanced regulation supports neural resilience and recovery processes. Thus, this datapoint serves as a key molecular indicator linking developmental neurobiology to psychiatric symptomatology, highlighting the importance of cellular differentiation mechanisms in both the etiology and treatment of mental disorders. |
| **Genetics and Molecular Biology:**<br><br>response to stress | The datapoint is classified in the Meta Mesh Ontology under NaturalProcess because it designates a temporally extended biological phenomenon in which an organism detects, interprets, and reacts to internal or external stressors through coordinated molecular, cellular, or systemic mechanisms. Within the BFO/MMO hierarchy, this aligns with entity → occurent unit → process unit → NaturalProcess, as the response is not a static continuant but a dynamic, causally | It encompasses the cellular, molecular, and systemic mechanisms by which the body adapts to internal or external stressors, processes that are directly implicated in the development and course of mental disorders. Clinically, dysregulation of stress-response pathways, including hypothalamic-pituitary-adrenal axis activity, inflammatory signaling, and neuroendocrine modulation, has been linked to depression, anxiety disorders, post-traumatic stress disorder, and stress-related somatic syndromes, making this biological process critical for both diagnosis and treatment. |

| | | |
|---|---|---|
| | structured unfolding that occurs across time. It cannot be categorized as a disposition, because it does not denote a latent capacity or inherent potential residing in the organism, but rather the realized enactment of biological adjustments to stress. Nor is it adequately modeled as a quality, since it is not a persisting attribute of an entity but an eventive and context-dependent sequence of interactions. By classifying it as a NaturalProcess, the ontology preserves coherence by situating the stress response as a concrete biological occurrence that is instantiated in specific contexts and unfolds in a temporally structured manner, consistent with the treatment of adaptive and regulatory pathways in living systems. | The presence of maladaptive or exaggerated stress responses provides interpretive value by signaling heightened vulnerability to psychiatric morbidity, impaired resilience, and increased risk of chronic symptom persistence, whereas balanced or normalized responses may indicate adaptive coping and therapeutic progress. Thus, this datapoint functions as a fundamental molecular marker bridging environmental adversity with psychiatric symptomatology, informing integrative assessment and guiding interventions aimed at restoring regulatory balance in stress-responsive systems. |
| **Genetics and Molecular Biology:**<br><br>regulation of cytosolic calcium ion concentration | The datapoint is classified in the Meta Mesh Ontology under NaturalProcess because it describes a temporally extended biological phenomenon in which cellular mechanisms actively control the intracellular levels of calcium ions. Within the BFO/MMO hierarchy, this aligns with entity → occurent unit → process unit → NaturalProcess, as the regulatory activity unfolds in time through molecular interactions and signaling cascades, rather than persisting as a static continuant. It cannot be categorized as a disposition, because it does not denote a latent capacity of a cell or organelle to regulate calcium but instead captures the realized occurrence of regulation as it happens. Nor is it adequately modeled as a quality, since it does not represent a persisting property of cytosol or ions but the dynamic adjustments of concentration levels. By situating it as a NaturalProcess, the ontology provides ontological precision by acknowledging this regulatory activity as an eventive physiological process intrinsic to cellular homeostasis, rather than as a static attribute or unrealized potential. | Calcium signaling serves as a fundamental mediator of neuronal excitability, synaptic plasticity, and neurotransmitter release, processes central to cognition, emotion, and behavior. Clinically, disturbances in calcium homeostasis have been implicated in the pathophysiology of mood disorders, schizophrenia, bipolar disorder, and neurodegenerative conditions, highlighting its role as both a vulnerability factor and a therapeutic target. The presence of dysregulated cytosolic calcium regulation provides interpretive value by indicating impaired synaptic function, altered neuroplasticity, and increased susceptibility to stress-related and affective dysregulation, whereas restoration of calcium balance may correspond with improved cognitive performance, mood stabilization, and treatment responsiveness. Thus, this datapoint functions as a critical molecular marker linking cellular signaling dynamics to psychiatric symptomatology, underscoring the need to integrate neurobiological mechanisms of calcium regulation into comprehensive psychiatric assessment and intervention strategies. |
| **Physiology and Biomarkers:**<br><br>Cortisol^AM peak specimen:MCnc:Pt:Ser/Plas:Qn | The datapoint is classified in the Meta Mesh Ontology under PhysiologicalQuality because it denotes a measurable, persisting feature of a biological system that inheres in serum or plasma as a medium of expression of endocrine regulation. Within the BFO/MMO hierarchy, this aligns with entity → continuant unit → sdc unit → quality unit → PhysiologicalQuality, since the cortisol concentration at a morning peak represents an | The morning cortisol peak reflects the functional integrity of the hypothalamic-pituitary-adrenal (HPA) axis, a central regulator of stress response and circadian rhythm. Clinically, deviations in the amplitude or timing of this peak are closely associated with major depressive disorder, anxiety disorders, post-traumatic stress disorder, and stress-related somatic syndromes, where altered HPA-axis activity serves as both a diagnostic biomarker and a predictor of treatment response. The presence of an attenuated or blunted morning cortisol |

| | | |
|---|---|---|
| | intrinsic physiological state rather than an independent object or a transient event. It cannot be modeled as a disposition, because it does not refer to a potential or capacity of the adrenal system, but rather to the actually instantiated level of hormone at a specific temporal point. Nor is it appropriately considered a process, since the datapoint does not describe the dynamic secretion or metabolic pathways of cortisol but the quantifiable state of concentration persisting in a biological specimen. By classifying it as a PhysiologicalQuality, the ontology preserves ontological rigor by situating the datapoint as a quality inherent in bodily systems, which can be quantitatively measured and compared across contexts while remaining distinct from latent capacities or unfolding processes. | rise may indicate chronic stress adaptation, burnout, or vulnerability to depressive relapse, while exaggerated peaks may signal heightened stress reactivity and increased risk of anxiety or insomnia. Thus, this datapoint provides interpretive value as a quantifiable physiological marker linking endocrine regulation to psychiatric symptomatology, enabling more precise assessment of stress-related pathology and guiding targeted interventions in psychiatric care. |
| **Physiology and Biomarkers:**<br><br>Heart rate:NRat:Pt:XXX:Qn | The datapoint is classified in the Meta Mesh Ontology under PhysiologicalQuality because it denotes a measurable, intrinsic feature of an organism's cardiovascular system that persists as a state and can be quantified at a given point in time. In the BFO/MMO hierarchy this corresponds to entity → continuant unit → sdc unit → quality unit → PhysiologicalQuality, since heart rate reflects a persisting physiological condition rather than a material entity or an unfolding occurrent. It is not appropriately modeled as a disposition, because it does not capture a latent capacity of the heart to beat but the realized, measurable state of its rhythm at the point of observation. Nor should it be treated as a process, since the term does not refer to the dynamic contraction of cardiac muscle fibers over time but rather to the quantifiable quality of beat frequency abstracted from that underlying activity. By situating it as a PhysiologicalQuality, the ontology provides a precise and consistent representation of heart rate as a quality that inheres in the organism and is accessible through biomarker measurement, thus distinguishing it from both unrealized potentials and temporally extended processes. | Autonomic regulation of cardiac activity is closely linked to emotional processing, arousal, and stress responsiveness. Clinically, alterations in resting heart rate or its variability are significant in the assessment of anxiety disorders, post-traumatic stress disorder, depression, and somatic symptom disorders, where autonomic dysregulation reflects underlying disturbances in stress physiology and emotional regulation. The presence of persistently elevated heart rate may indicate heightened sympathetic arousal, hypervigilance, or chronic anxiety, while reduced or blunted variability can reflect impaired parasympathetic control, stress exhaustion, or depressive states. Conversely, balanced and adaptive heart rate regulation is associated with resilience, emotional stability, and recovery. Thus, this datapoint provides interpretive value as a readily measurable physiological marker that bridges autonomic function with psychiatric symptomatology, supporting both diagnostic assessment and monitoring of treatment outcomes in mental health care. |
| **Physiology and Biomarkers:**<br><br>Body mass index:Ratio:Pt:^P | The datapoint is classified in the Meta Mesh Ontology under PhysiologicalRelationalQuality because it denotes a quantitative quality that arises from the relation between two physiological measures, namely body mass and height, rather than a simple intrinsic attribute of an anatomical | Weight regulation and metabolic status are closely intertwined with mental health through biological, behavioral, and psychosocial pathways. Clinically, abnormal BMI values are important for assessing risk in eating disorders, depression, bipolar disorder, and schizophrenia, where both underweight and obesity may reflect underlying |

| | | |
|---|---|---|
| atient:Qn | structure. Within the BFO/MMO hierarchy, this corresponds to entity → continuant unit → sdc unit → quality unit → relational quality unit → PhysiologicalRelationalQuality, since body mass index inheres in the organism as a persisting, measurable state but is constituted through a ratio that relates otherwise independent physical dimensions of the same body. It cannot be categorized as a disposition, because it does not represent a potential or latent capacity of the organism, nor is it a process, since it does not describe a dynamic event of growth, metabolism, or physical activity unfolding in time. Instead, BMI captures a relatively stable, relationally defined physiological quality that persists independently of immediate occurrent activity and provides a structured measure of body composition. By situating it as a PhysiologicalRelationalQuality, the ontology ensures ontological precision by recognizing its dependence on the comparative relation of mass to height while maintaining its status as an inhering, quantifiable quality of the patient's body. | psychopathology as well as the metabolic side effects of psychotropic medication. The presence of significantly low BMI can indicate restrictive eating patterns, malnutrition, or severe depressive states, while elevated BMI may reveal emotional dysregulation, stress-related eating, or antipsychotic-induced metabolic disturbances. Stable and normative BMI, by contrast, may suggest balanced lifestyle patterns and treatment stability. Thus, this datapoint provides interpretive value as an accessible and quantifiable marker that connects somatic health with psychiatric status, informing both diagnostic evaluation and the monitoring of therapeutic outcomes in mental health care. |
| **Physiology and Biomarkers:**<br><br>Calcidiol+ercalcidiol:MCnc:Pt:Ser/Plas:Qn | The datapoint is classified in the Meta Mesh Ontology under PhysiologicalQuality because it designates a quantifiable concentration of vitamin D metabolites that inheres in a biological specimen as a persisting state of the organism's physiology. In the BFO/MMO hierarchy, this corresponds to entity → continuant unit → sdc unit → quality unit → PhysiologicalQuality, since the measurement reflects an intrinsic property of the serum or plasma, describing the amount of calcidiol and ercalcidiol present at a given time. It is not properly understood as a disposition, because the datapoint does not describe a potential of the organism to metabolize vitamin D but instead the actualized state of metabolite concentration. Nor should it be modeled as a process, since the term does not capture the dynamic biochemical pathways that produce or consume these metabolites but the measurable quality that results from them. By classifying it as a PhysiologicalQuality, the ontology ensures ontological precision by situating this biomarker as a stable, inhering quality of the organism that is susceptible to quantitative assessment while remaining distinct from capacities or event-like | Serum levels of vitamin D metabolites play a critical role in neuroendocrine regulation, neuroplasticity, and immune modulation, all of which influence mental health outcomes. Clinically, insufficient concentrations of calcidiol and ercalcidiol have been associated with increased risk of depressive disorders, seasonal affective disorder, cognitive decline, and greater symptom severity in schizophrenia and bipolar disorder, underscoring their importance in both assessment and treatment planning. The presence of deficiency may signal impaired neurobiological resilience, heightened vulnerability to mood dysregulation, and reduced capacity for stress adaptation, while sufficient or normalized levels can support improved affective stability and cognitive functioning. Thus, this datapoint provides interpretive value as a measurable biomarker linking metabolic and nutritional status to psychiatric symptomatology, informing integrative care strategies that combine psychopharmacological, psychosocial, and metabolic interventions. |

| | | |
|---|---|---|
| | processes. | |
| **Physiology and Biomarkers:**<br><br>C reactive protein:MCnc:Pt:Ser/Plas:Qn | The datapoint is classified in the Meta Mesh Ontology under PhysiologicalQuality because it designates a measurable concentration of a biomolecule that inheres in a biological specimen as an intrinsic feature of the organism's physiological state. Within the BFO/MMO hierarchy, this corresponds to entity → continuant unit → sdc unit → quality unit → PhysiologicalQuality, since the concentration of C reactive protein is a persisting, quantifiable attribute of the serum or plasma at a given point in time rather than an independent object or a temporally extended event. It cannot be appropriately modeled as a disposition, because the datapoint does not describe the potential of the liver to synthesize acute-phase proteins but the actually realized concentration of CRP present in the bloodstream. Nor is it a process, since it does not denote the dynamic biochemical pathways of protein production or clearance but instead the static, measurable level that results from those processes. By situating the datapoint as a PhysiologicalQuality, the ontology ensures precise alignment with BFO's continuant framework, capturing CRP concentration as an inhering and quantifiable quality of the organism's physiological condition. | Elevated CRP levels serve as a peripheral marker of systemic inflammation, a biological process increasingly recognized as a contributor to the pathophysiology of mental disorders. Clinically, heightened CRP concentrations have been associated with major depressive disorder, bipolar disorder, schizophrenia, and post-traumatic stress disorder, where inflammatory dysregulation may exacerbate symptom severity, treatment resistance, and cognitive impairment. The presence of elevated CRP provides interpretive value by indicating an inflammatory state that may underlie or aggravate psychiatric symptomatology, suggesting the need for integrative treatment approaches targeting both inflammation and mental health, while low or normalized CRP levels may reflect reduced biological stress burden and improved clinical stability. Thus, this datapoint functions as a measurable biomarker linking immune activity to psychiatric outcomes, supporting more precise assessment and personalized intervention strategies within psychiatric care. |
| **Behavioral and Lifestyle Data:**<br><br>Detachment | The datapoint is classified in the Meta Mesh Ontology under PsychologicalDisposition because it designates a stable, person-centered tendency of psychological functioning that inheres in an individual across time and contexts. Within the BFO/MMO hierarchy, it is situated under entity → continuant unit → sdc unit → realizable unit → disposition unit → PersonCenteredDisposition → PsychologicalDisposition, as it represents a realizable quality that manifests in behavioral and experiential patterns but is not itself a transient event. Unlike a process, which unfolds as a temporally extended occurrence such as a social interaction or cognitive episode, detachment persists as an underlying capacity or orientation that can be expressed in multiple processes without being reducible to them. Nor does it correspond to a mere quality, since it does not denote a static measurable state but rather a structured potential for affective and interpersonal disengagement. By | It reflects a persistent pattern of emotional and interpersonal disengagement that directly influences social functioning, motivation, and well-being. Clinically, detachment is a central construct in the dimensional assessment of personality pathology, depressive syndromes, and trauma-related disorders, where it captures difficulties in experiencing positive affect, forming close relationships, and engaging with the external world. The presence and severity of detachment provide interpretive value by indicating reduced capacity for social connectedness, heightened vulnerability to loneliness and anhedonia, and impaired treatment engagement, while its absence or attenuation may signal greater resilience, improved affective responsiveness, and more adaptive interpersonal functioning. Thus, this datapoint functions as a clinically meaningful marker of socioemotional withdrawal that informs diagnostic formulation, prognostic evaluation, and therapeutic strategies in psychiatric care. |

| | | |
|---|---|---|
| | classifying it as a PsychologicalDisposition, the MMO preserves the ontological distinction between enduring dispositions that condition behavior and the processes in which those dispositions are episodically realized. | |
| **Behavioral and Lifestyle Data:**<br><br>Disinhibition | The datapoint is classified in the Meta Mesh Ontology under PsychologicalDisposition because it represents a stable, person-centered tendency of mental and behavioral regulation that inheres in the individual as a realizable continuant. Within the BFO/MMO hierarchy, it is situated under entity → continuant unit → sdc unit → realizable unit → disposition unit → PersonCenteredDisposition → PsychologicalDisposition, since it refers to an enduring predisposition that structures how an individual modulates impulses and responses across different contexts. It is not to be understood as a process, since it does not describe a temporally extended sequence of inhibitory failures or impulsive actions but rather the underlying dispositional ground from which such processes may emerge. Nor does it qualify as a mere quality, as disinhibition is not a static, measurable state at a point in time but a structured potential for characteristic patterns of affective and behavioral expression. By situating it as a PsychologicalDisposition, the MMO ensures an ontologically precise treatment that distinguishes the enduring propensity from the episodic processes in which it is manifested. | It denotes a tendency toward impulsivity, poor self-regulation, and diminished capacity to restrain inappropriate behaviors or emotions, features that are strongly implicated in the onset and maintenance of psychopathology. Clinically, the assessment of disinhibition is critical in disorders such as attention-deficit/hyperactivity disorder, substance use disorders, and certain personality disorders, where impaired inhibitory control contributes to functional impairment, risky behavior, and treatment nonadherence. The presence and severity of disinhibition provide interpretive value by signaling heightened vulnerability to maladaptive decision-making, emotional dysregulation, and externalizing pathology, while its absence or reduction may reflect improved executive functioning, affective stability, and therapeutic progress. Thus, this datapoint serves as a key behavioral marker that bridges neurocognitive mechanisms of control with psychiatric symptomatology, supporting diagnostic clarity and informing interventions that target self-regulation and impulse control. |
| **Behavioral and Lifestyle Data:**<br><br>Avolition | The datapoint is classified in the Meta Mesh Ontology under PsychologicalDisposition because it denotes an enduring tendency of diminished motivation and goal-directed behavior that inheres in the individual as a stable realizable attribute. Within the BFO/MMO hierarchy, it is situated under entity → continuant unit → sdc unit → realizable unit → disposition unit → PersonCenteredDisposition → PsychologicalDisposition, since it describes a persisting potential or inclination that conditions how an individual engages with activities over time. It does not correspond to a process, as it does not refer to the unfolding of a specific episode of inactivity or reduced initiation of behavior, but rather to the dispositional basis from which such processes emerge. Nor can it be adequately modeled as a static quality, because avolition is not | It reflects a marked reduction in the initiation and persistence of goal-directed activity, a core symptom domain that profoundly impacts functional capacity and quality of life. Clinically, avolition is especially salient in schizophrenia, major depressive disorder, and other severe mental illnesses, where diminished motivation and drive contribute to social withdrawal, occupational decline, and poor treatment adherence. The presence and severity of avolition provide interpretive value by signaling underlying deficits in reward processing, executive functioning, and volitional capacity, while its absence or improvement may indicate therapeutic progress, restored motivation, and enhanced psychosocial functioning. Thus, this datapoint functions as a critical indicator of negative symptomatology and functional impairment, informing diagnostic assessment, prognosis, and the design of interventions aimed at restoring motivational and behavioral engagement in psychiatric care. |

| | | |
|---|---|---|
| | a momentary measurable property but an enduring predisposition that shapes the likelihood of motivational deficits being expressed across contexts. By classifying it as a PsychologicalDisposition, the MMO ensures ontological precision by distinguishing avolition as an underlying dispositional orientation, differentiating it from transient processes or superficial qualities, and thereby preserving its status as a person-centered psychological trait. | |
| **Behavioral and Lifestyle Data:**<br><br>Anhedonia | The datapoint is classified in the Meta Mesh Ontology under PsychologicalDisposition because it denotes a persistent tendency of diminished capacity to experience pleasure or interest that inheres in the individual as a realizable continuant. Within the BFO/MMO hierarchy, it is positioned under entity → continuant unit → sdc unit → realizable unit → disposition unit → PersonCenteredDisposition → PsychologicalDisposition, since it describes a stable, person-centered predisposition that structures the likelihood of specific affective and behavioral responses rather than a transient occurrence. It is not adequately characterized as a process, as it does not describe the unfolding of particular episodes of diminished pleasure but instead the underlying predisposition that explains why such episodes recur across contexts. Nor is it properly modeled as a static quality, because anhedonia is not a fixed measurable state at a point in time but an enduring dispositional orientation shaping experiential potential. By classifying it as a PsychologicalDisposition, the MMO maintains ontological precision by situating anhedonia as a persistent predispositional attribute that grounds, but is distinct from, the transient processes in which it is expressed. | It represents a diminished capacity to experience pleasure or interest, reflecting fundamental disruptions in reward processing and affective responsiveness. Clinically, anhedonia is a hallmark symptom of major depressive disorder and a critical feature in schizophrenia, post-traumatic stress disorder, and substance use disorders, where it serves as a key marker of illness severity, chronicity, and treatment resistance. The presence and intensity of anhedonia provide interpretive value by indicating impaired motivational drive, reduced engagement with rewarding activities, and elevated risk of functional decline or suicidality, while its absence or remission may signal therapeutic efficacy and improved quality of life. Thus, this datapoint functions as a clinically and prognostically significant indicator that bridges neurobiological reward circuitry with observable behavior, supporting precise psychiatric assessment and the tailoring of interventions aimed at restoring hedonic capacity and social functioning. |
| **Behavioral and Lifestyle Data:**<br><br>Antagonism | The datapoint is classified in the Meta Mesh Ontology under PsychologicalDisposition because it represents an enduring person-centered predisposition toward hostile, oppositional, or conflict-prone patterns of thought and behavior. Within the BFO/MMO hierarchy, it is positioned under entity → continuant unit → sdc unit → realizable unit → disposition unit → PersonCenteredDisposition → PsychologicalDisposition, as it denotes a realizable continuant that inheres in the individual and is | It reflects a persistent pattern of hostility, oppositionality, and insensitivity to the needs or rights of others, traits that disrupt social functioning and are closely tied to externalizing psychopathology. Clinically, the assessment of antagonism is essential in the evaluation of personality disorders, conduct disorder, substance use disorders, and certain mood and impulse-control conditions, where antagonistic traits contribute to interpersonal conflict, aggression, and poor treatment engagement. The presence and severity of antagonism provide interpretive value by signaling heightened risk of maladaptive relational patterns, |

| | | |
|---|---|---|
| | manifested in repeated interpersonal contexts. It is not adequately described as a process, since antagonistic episodes are temporally extended events that only instantiate the underlying tendency but do not exhaust it. Nor can it be reduced to a mere quality, because antagonism is not a static, momentary property but an enduring structured potential shaping affective and behavioral regulation. By situating it as a PsychologicalDisposition, the MMO captures its ontological status as a stable predispositional orientation that conditions the likelihood of antagonistic behaviors and experiences while remaining distinct from the transient processes in which it is expressed. | reduced prosocial behavior, and increased likelihood of comorbidity with antisocial or narcissistic features, while its absence or attenuation may indicate improved empathy, cooperation, and therapeutic alliance. Thus, this datapoint serves as a clinically meaningful marker that links interpersonal dysfunction to psychiatric morbidity, informing diagnostic formulation, prognostic evaluation, and the tailoring of interventions aimed at improving relational and behavioral regulation. |
| **Neurocognitive and Psychometric Data:**<br><br>Digit span reverse | The datapoint is classified in the Meta Mesh Ontology under PsychologicalDisposition because it denotes an enduring capacity of an individual to temporarily store and manipulate information in working memory. Within the BFO/MMO hierarchy, this entity is situated under entity → continuant unit → sdc unit → realizable unit → disposition unit → PersonCenteredDisposition → PsychologicalDisposition, since it represents a realizable continuant inhering in the subject that is manifested when cognitive demands elicit the ability to recall and reverse sequences of digits. It is not reducible to a mere quality, as it is not a static property observable at a single time point, nor is it a process, since the cognitive testing event is only the actualization of the underlying ability and not identical with it. By classifying "Digit span reverse" as a PsychologicalDisposition, the MMO captures its ontological status as a stable, person-centered potential for cognitive performance that persists across time and contexts, grounding the observable outcomes of specific psychometric assessments in a structured dispositional framework. | It directly assesses working memory, attentional control, and executive functioning, cognitive domains that are frequently impaired across a wide range of mental disorders. Clinically, performance on this task is particularly important in the evaluation of schizophrenia, major depressive disorder, bipolar disorder, attention-deficit/hyperactivity disorder, and neurocognitive disorders, where deficits in working memory contribute to functional impairment, poor treatment adherence, and reduced social and occupational capacity. The presence of diminished digit span reverse performance provides interpretive value by indicating compromised prefrontal cortical functioning, heightened vulnerability to cognitive overload, and difficulties with complex task execution, while preserved or improved performance may signal cognitive resilience, therapeutic efficacy, or recovery of executive functioning. Thus, this datapoint serves as a sensitive cognitive marker linking neuropsychological processes to psychiatric symptomatology, supporting diagnostic assessment, prognosis, and the development of interventions targeting cognitive remediation. |
| **Neurocognitive and Psychometric Data:**<br><br>California verbal learning test | The datapoint is classified in the Meta Mesh Ontology under PsychologicalDisposition because it refers not to the testing procedure itself but to the enduring cognitive capacity that the test is designed to elicit and measure, namely verbal learning and memory. Within the BFO/MMO hierarchy, it is appropriately situated under entity → continuant unit → sdc unit → realizable unit → disposition unit → PersonCenteredDisposition → | It provides a standardized measure of verbal learning, memory encoding, retrieval, and recognition, domains frequently impaired in psychiatric and neurocognitive disorders. Clinically, this test is particularly valuable in the assessment of schizophrenia, major depressive disorder, bipolar disorder, post-traumatic stress disorder, and dementia syndromes, where deficits in verbal memory serve as both diagnostic indicators and predictors of functional outcome. The presence of impaired performance on the |

| | | |
|---|---|---|
| | PsychologicalDisposition, since it denotes a realizable continuant that inheres in the subject as a latent potential, which is manifested when the individual engages in structured recall and learning tasks. It is not reducible to a process, as the act of administering the test constitutes a temporally extended event, whereas the underlying capacity measured by it persists independently of any particular testing session. Nor is it a static quality, because the ability to encode and retrieve verbal information is not an instantaneous observable property but a stable predisposition that grounds the occurrence of successful or impaired learning episodes. By classifying it as a PsychologicalDisposition, the MMO ensures ontological precision by recognizing the California verbal learning test as an instrumentally mediated reference to an enduring person-centered capacity, thereby distinguishing the dispositional basis from its empirical assessment process. | California Verbal Learning Test provides interpretive value by signaling disruptions in hippocampal and prefrontal cortical functioning, highlighting vulnerability to cognitive decline, and revealing the extent of impairment that may compromise treatment adherence and daily functioning, while intact or improved performance may reflect cognitive resilience, therapeutic benefit, or recovery of mnemonic processes. Thus, this datapoint functions as a sensitive neuropsychological marker linking cognitive mechanisms to psychiatric symptomatology, informing diagnosis, prognosis, and the development of individualized cognitive and clinical interventions. |
| **Neurocognitive and Psychometric Data:**<br><br>Stroop Test | The datapoint is classified in the Meta Mesh Ontology under PsychologicalDisposition because it designates not the testing procedure itself but the enduring cognitive capacities that the procedure is intended to assess, namely selective attention, cognitive control, and interference inhibition. Within the BFO/MMO hierarchy, it is appropriately placed under entity → continuant unit → sdc unit → realizable unit → disposition unit → PersonCenteredDisposition → PsychologicalDisposition, since it refers to a realizable continuant that inheres in the individual and is manifested when the subject engages in tasks requiring inhibition of automatic responses and flexible allocation of attention. It is not reducible to a process, because the administration of the Stroop Test is a temporally bounded assessment event, whereas the underlying inhibitory control and attentional regulation persist independently as dispositional potentials. Nor can it be modeled as a static quality, because these cognitive traits are not momentary measurable states but structured predispositions that ground a range of observable performances across contexts. By situating it as a PsychologicalDisposition, the MMO distinguishes the enduring cognitive capacity that the Stroop Test measures from the test procedure itself, thereby preserving | It measures cognitive control, selective attention, and response inhibition, functions that are frequently disrupted across a spectrum of psychiatric disorders. Clinically, performance on the Stroop Test is particularly informative in conditions such as schizophrenia, major depressive disorder, bipolar disorder, attention-deficit/hyperactivity disorder, and substance use disorders, where deficits in executive functioning contribute to impaired decision-making, emotional dysregulation, and reduced adaptive capacity. The presence of Stroop interference effects, reflected in slowed response times or increased errors, provides interpretive value by indicating diminished prefrontal cortical regulation and heightened susceptibility to cognitive intrusions, while preserved or improved performance may reflect effective compensatory mechanisms, treatment responsiveness, or cognitive resilience. Thus, this datapoint functions as a sensitive and widely validated neuropsychological marker linking executive dysfunction to psychiatric symptomatology, guiding diagnostic assessment, prognostic evaluation, and the design of targeted cognitive and therapeutic interventions. |

| | | |
|---|---|---|
| | ontological clarity between the dispositional substrate and its empirical elicitation. | |
| **Neurocognitive and Psychometric Data:**<br><br>TMT part A | The datapoint is classified in the Meta Mesh Ontology under PsychologicalDisposition because it operationalizes an enduring cognitive capacity rather than a transient event. Within the BFO/MMO hierarchy, it is positioned under entity → continuant unit → sdc unit → realizable unit → disposition unit → PersonCenteredDisposition → PsychologicalDisposition, as it refers to the stable cognitive disposition of visual attention, processing speed, and sequencing ability that the Trail Making Test part A is designed to measure. Although the test itself is administered as a process, the underlying target of measurement- the individual's dispositional capacity to efficiently sustain attention and organize visuomotor responses - exists independently of any single test performance and persists across contexts. It cannot be reduced to a mere process, since the administration of the task is an event that reveals, rather than constitutes, the disposition. Nor is it adequately captured as a static quality, as the construct of attention and processing speed reflects a realizable potential that manifests variably depending on situational demands. Classifying the datapoint as a PsychologicalDisposition therefore maintains ontological consistency by distinguishing the enduring cognitive potential being measured from its empirical elicitation in a specific testing procedure. | It assesses processing speed, visual attention, and psychomotor efficiency, cognitive domains that are frequently affected in mental disorders. Clinically, performance on this test is particularly informative in depression, schizophrenia, bipolar disorder, and neurocognitive conditions, where slowed processing speed and attentional deficits are core features contributing to functional impairment and poor treatment adherence. The presence of prolonged completion time or elevated error rates on TMT part A provides interpretive value by indicating cognitive slowing, reduced attentional capacity, or psychomotor retardation, while normal or improved performance may suggest preserved cognitive efficiency, resilience, or positive treatment response. Thus, this datapoint functions as a sensitive and practical neuropsychological marker that links basic cognitive processing to psychiatric symptomatology, supporting diagnostic evaluation, prognosis, and the design of interventions aimed at enhancing cognitive and functional outcomes. |
| **Neurocognitive and Psychometric Data:**<br><br>Wisconsin Card Sorting Test | The datapoint is classified in the Meta Mesh Ontology under PsychologicalDisposition because it designates the enduring cognitive capacities of cognitive flexibility, rule-shifting, and executive control that the test is designed to elicit and measure. Within the BFO/MMO hierarchy, it is situated under entity → continuant unit → sdc unit → realizable unit → disposition unit → PersonCenteredDisposition → PsychologicalDisposition, since it refers to a realizable continuant that inheres in the subject as a latent potential to adapt cognitive strategies in response to changing environmental contingencies. It is not a process, as the administration of the Wisconsin Card Sorting Test constitutes a bounded event of assessment, while the underlying capacities it measures persist independently of any | It measures executive functions such as cognitive flexibility, abstract reasoning, and set-shifting, which are central to adaptive problem-solving and behavioral regulation. Clinically, impaired performance on this test is particularly significant in schizophrenia, bipolar disorder, major depressive disorder, and neurocognitive disorders, where deficits in executive control contribute to poor functional outcomes, impaired insight, and difficulties in treatment adherence. The presence of perseverative errors or reduced ability to shift strategies provides interpretive value by indicating prefrontal cortical dysfunction, rigidity in thought processes, and vulnerability to maladaptive coping, while preserved or improved performance may reflect cognitive resilience, therapeutic responsiveness, or intact executive capacity. Thus, this datapoint functions as a critical neuropsychological marker linking executive dysfunction to psychiatric symptomatology, informing diagnostic |

| | | |
|---|---|---|
| | specific testing episode. Nor is it a static quality, because cognitive flexibility is not reducible to an instantaneous observable property but represents an enduring predisposition structuring how an individual can respond across diverse contexts. By classifying it as a PsychologicalDisposition, the MMO preserves ontological clarity by recognizing the Wisconsin Card Sorting Test as an empirical probe of an underlying dispositional capacity, thereby separating the measurement procedure from the enduring psychological potential it reveals. | formulation, prognosis, and the development of interventions that target cognitive remediation and functional rehabilitation. |
| **Sensor Data:**<br><br>Night-time screen activity | The datapoint is classified in the Meta Mesh Ontology under TechnicalProcess because it denotes a temporally extended sequence of operations executed by a technical device, rather than a static attribute or enduring capacity. Within the BFO/MMO hierarchy, it falls under entity → occurent unit → process unit → TechnicalProcess, since the activity consists of the screen being switched on, illuminated, and interacted with over time, reflecting a series of technical state changes and user-device interactions. It is not adequately modeled as a disposition, because the datapoint does not refer to a latent potential of the device but instead captures an actualized event unfolding in time. Nor is it properly classified as a quality, since it does not describe a static property of the device but rather a dynamic occurrence detectable through sensor monitoring. Classifying this datapoint as a TechnicalProcess ensures ontological consistency, as it captures the temporal, processual nature of the screen's operational behavior during night-time intervals. | Late-hour digital engagement disrupts circadian rhythms, impairs sleep quality, and is strongly associated with affective and cognitive dysregulation. Clinically, monitoring screen use during nocturnal hours provides valuable insight into the behavioral patterns of individuals with insomnia, depression, bipolar disorder, anxiety disorders, and attention-deficit/hyperactivity disorder, where disturbed sleep-wake cycles and compulsive technology use frequently exacerbate symptom severity. The presence and intensity of night-time screen activity provide interpretive value by signaling heightened arousal, poor self-regulation, and vulnerability to mood instability, while its absence or reduction may indicate healthier sleep hygiene, restored circadian stability, and improved psychiatric functioning. Thus, this datapoint functions as an ecologically valid behavioral marker linking digital behavior and circadian disruption to psychiatric symptomatology, supporting both diagnostic assessment and the tailoring of interventions aimed at improving sleep, emotional regulation, and overall mental health. |
| **Sensor Data:**<br><br>Circadian activity pattern | The datapoint is classified in the Meta Mesh Ontology under NaturalProcess because it captures a temporally extended, recurrent biological rhythm manifested through observable fluctuations in activity levels. Within the BFO/MMO hierarchy, it is situated under entity → occurent unit → Process unit → NaturalProcess, as it denotes an ongoing physiological process rooted in endogenous circadian regulation and expressed through locomotor activity across day-night cycles. It cannot be adequately modeled as a disposition, since the circadian rhythm is not merely a latent potential but an actualized, temporally unfolding process that continually structures behavior. Nor | Daily rhythms of physical activity serve as objective markers of sleep-wake regulation, behavioral consistency, and overall mental health. Clinically, disrupted circadian activity is strongly associated with major depressive disorder, bipolar disorder, schizophrenia, and anxiety disorders, where irregularities in rest-activity cycles exacerbate symptom burden and impair treatment response. The presence of fragmented or delayed circadian patterns provides interpretive value by indicating dysregulated biological rhythms, diminished behavioral activation, and elevated vulnerability to mood instability, while regular and stable activity patterns may reflect resilience, improved self-regulation, and therapeutic progress. Thus, this datapoint functions as a reliable, ecologically valid |

| | | |
|---|---|---|
| | does it qualify as a static quality, as it is defined by rhythmic variation and change over time rather than by an instantaneous or enduring property. By classifying it as a NaturalProcess, the MMO ensures ontological consistency, representing circadian activity as a processual phenomenon embedded in biological timekeeping rather than as a capacity or a mere measurement. | biomarker linking motor activity and circadian organization to psychiatric symptomatology, supporting more precise assessment and informing interventions aimed at stabilizing daily rhythms to promote psychiatric recovery. |
| **Sensor Data:**<br><br>Typing behavior | The datapoint is classified in the Meta Mesh Ontology under TechnicalProcess because it denotes a temporally extended sequence of operations occurring through the interaction between a user and a technical device. Within the BFO/MMO hierarchy, it is situated under entity → occurent unit → Process unit → TechnicalProcess, since typing behavior consists of discrete but ordered events - key presses, input recognition, and system-level responses - that unfold dynamically in time and can be continuously monitored by sensors. It is not adequately captured as a disposition, because the datapoint refers not to a latent potential for interaction but to the actualized event structure of interaction itself. Nor can it be modeled as a static quality, as typing behavior is defined by its temporal unfolding, variability, and rhythm rather than by a fixed property of either the user or the device. By classifying it as a TechnicalProcess, the MMO preserves ontological rigor by recognizing that what is being recorded is the technical event of keystroke activity as it occurs, not merely a capacity or abstract characteristic. | Fine-grained patterns of digital interaction provide ecologically valid insights into cognitive processing, psychomotor activity, and affective states. Clinically, alterations in keystroke dynamics, such as typing speed, latency, error rates, or rhythm, have been linked to depressive episodes, bipolar mood fluctuations, anxiety, and neurocognitive decline, offering potential for unobtrusive monitoring of symptom trajectories and treatment response. The presence of slowed or erratic typing patterns may indicate psychomotor retardation, attentional impairment, or mood-related cognitive load, while preserved fluency and consistency may reflect cognitive stability and therapeutic progress. Thus, this datapoint functions as a non-invasive digital biomarker that connects everyday behavioral expression with underlying psychiatric states, enabling more precise assessment, real-time monitoring, and individualized intervention strategies in mental health care. |
| **Sensor Data:**<br><br>Communication behavior | The datapoint is classified in the Meta Mesh Ontology under SocialProcess because it denotes a temporally extended interaction between persons mediated by communicative acts such as phone calls. Within the BFO/MMO hierarchy, this classification places it under entity → occurent unit → process unit → SocialProcess, since what is observed is the unfolding of social interaction across time, consisting of acts of initiation, response, and exchange that are inherently relational and context-dependent. It cannot be understood as a disposition, because the datapoint does not denote a latent capacity to communicate but instead captures the realized sequence of social activity. Nor is it reducible to a static quality, as its defining features lie in the temporality and | The frequency, duration, and patterns of interpersonal contact captured through digital communication serve as ecologically valid indicators of social engagement and relational functioning. Clinically, alterations in communication behavior are highly informative in mood disorders, psychotic disorders, anxiety disorders, and personality pathology, where social withdrawal, heightened contact seeking, or erratic interaction patterns reflect core dimensions of psychopathology. The presence of reduced or absent communication may indicate social isolation, anhedonia, or depressive states, while excessive or disorganized calling patterns may suggest anxiety, mania, or impaired impulse control; conversely, stable and balanced communication activity may reflect psychosocial functioning and therapeutic progress. Thus, this datapoint functions as a dynamic behavioral marker linking everyday |

| | | |
|---|---|---|
| | interactivity of communicative exchanges, which may vary in duration, frequency, and structure. Classifying it as a SocialProcess preserves ontological precision by recognizing that what is being measured through the sensor system is the occurrence of socially embedded interactions as they take place, situating them within the broader domain of socially constituted processes. | social behavior to psychiatric symptomatology, supporting diagnostic assessment, longitudinal monitoring, and the development of personalized interventions that integrate clinical care with real-world behavioral patterns. |
| **Sensor Data:**<br><br>GPS-based mobility pattern | The datapoint is classified in the Meta Mesh Ontology under NaturalProcess because it represents the spatiotemporal unfolding of physical movement in the natural environment, as captured through geolocation data. Within the BFO/MMO hierarchy, this classification situates the entity under occurent unit → process unit → NaturalProcess, since mobility patterns denote ongoing dynamic changes in spatial position that unfold through time and are embedded in the natural domain of bodily locomotion and environmental interaction. This ontological placement is justified because the datapoint does not capture a static feature or quality of an individual, nor does it denote a mere latent capacity for movement, but rather documents the realized trajectory of movement as a temporally extended process. It is not a disposition, since it refers to observed occurrences rather than inherent capabilities, and it is not adequately represented as a social or technical process, as the defining characteristic lies in the natural continuity of physical motion independent of institutional, symbolic, or technical mediation. Thus, classifying GPS-based mobility pattern as a NaturalProcess secures conceptual accuracy by aligning the datapoint with its ontological nature as a realized, time-bound process of spatial locomotion. | Spatial movement and activity patterns provide objective markers of behavioral activation, social engagement, and circadian regulation, all of which are closely tied to mental health. Clinically, reduced mobility or restricted geographic range has been associated with depressive disorders, social withdrawal, and negative symptomatology in schizophrenia, while erratic or excessive mobility may indicate manic states, anxiety-driven avoidance, or impulsive behavior. The presence of diminished mobility patterns provides interpretive value by signaling functional decline, loss of motivation, or social isolation, whereas preserved or normalized mobility may reflect improved mood, treatment responsiveness, and greater integration into daily life. Thus, this datapoint functions as an ecologically valid behavioral biomarker that links real-world activity to psychiatric symptomatology, supporting diagnostic assessment, longitudinal monitoring, and the tailoring of interventions aimed at restoring functional and social participation. |
| **Previous Diagnoses:**<br><br>Social Phobias | The datapoint is classified in the Meta Mesh Ontology under PsychologicalDisposition because it denotes a persistent, person-centered tendency toward pathological fear responses in social contexts rather than an episodic occurrence or a static state. Within the BFO/MMO hierarchy, this classification is situated under continuant unit → sdc unit → realizable unit → disposition unit → PersonCenteredDisposition → PsychologicalDisposition, since social phobias describe an enduring psychological vulnerability that may manifest under particular circumstances | They reflect enduring patterns of excessive fear and avoidance of social or performance situations that significantly impair daily functioning and quality of life. Clinically, a history of social phobia is essential to consider in psychiatric assessment because it frequently co-occurs with depressive disorders, substance use disorders, and other anxiety conditions, and it often shapes the course of illness through chronicity, reduced social integration, and heightened vulnerability to isolation. The presence and severity of this diagnosis provide interpretive value by indicating longstanding difficulties in interpersonal engagement, elevated risk for secondary psychopathology, |

| | | |
|---|---|---|
| | but exists as a latent potential even in the absence of actual social interaction. This ontological placement distinguishes it from processes, which would capture the concrete unfolding of phobic reactions during specific social encounters, and from states, which would refer to a temporally bound observable condition such as acute anxiety at a given moment. By contrast, social phobias inherently describe a realizable disposition that structures the likelihood of fear and avoidance behaviors across time, making PsychologicalDisposition the most accurate and consistent classification. | and potential barriers to treatment adherence due to avoidance tendencies, while its absence or successful treatment may suggest improved psychosocial functioning, resilience, and therapeutic progress. Thus, this datapoint functions as a critical historical marker linking prior anxiety pathology to current psychiatric state, guiding both diagnostic formulation and the development of interventions that address residual social and functional impairments. |
| **Previous Diagnoses:**<br><br>Agoraphobia | The datapoint is classified in the Meta Mesh Ontology under PsychologicalDisposition because it designates a persistent psychological tendency that predisposes individuals to experience intense fear and avoidance in situations where escape may be perceived as difficult. Within the BFO/MMO hierarchy, this entity is correctly situated under continuant unit → sdc unit → realizable unit → disposition unit → PersonCenteredDisposition → PsychologicalDisposition, as agoraphobia is not an event that unfolds in time but rather an enduring dispositional structure that manifests under specific contextual triggers. It cannot be adequately modeled as a process, since the actual panic or avoidance episodes represent contingent realizations of the disposition rather than the disposition itself, and it is not a static quality, because its essence lies in the capacity to generate fear responses across varying circumstances rather than in a temporally fixed property. The classification as PsychologicalDisposition ensures ontological precision by capturing agoraphobia as a latent but enduring mental vulnerability that organizes the probability of certain behavioral and emotional outcomes, thereby preserving the distinction between realized episodes and the underlying predispositional basis. | It denotes a pervasive anxiety condition characterized by fear and avoidance of situations where escape might be difficult or help unavailable, leading to substantial restrictions in mobility and independence. Clinically, a history of agoraphobia is highly informative, as it frequently co-occurs with panic disorder, major depressive disorder, and substance misuse, and it often contributes to chronic disability, diminished quality of life, and increased healthcare utilization. The presence and severity of this diagnosis provide interpretive value by signaling persistent avoidance behavior, heightened vulnerability to social and occupational impairment, and elevated risk of psychiatric comorbidity, while its absence or remission may reflect successful treatment, improved coping strategies, and enhanced psychosocial functioning. Thus, this datapoint serves as an important clinical marker linking prior anxiety pathology to present psychiatric status, informing diagnostic formulation, prognosis, and the tailoring of therapeutic interventions aimed at restoring autonomy and reducing avoidance. |
| **Previous Diagnoses:**<br><br>Anankastic personality | The datapoint is classified in the Meta Mesh Ontology under PsychologicalDisposition because it refers to a persistent, person-centered predisposition that structures characteristic patterns of cognition, emotion, and behavior rather than an episodic event or a fixed state. Within the BFO/MMO hierarchy, this classification is situated | It reflects a pervasive pattern of perfectionism, rigidity, and excessive control that profoundly influences emotional regulation, interpersonal relationships, and functional capacity. Clinically, a history of this disorder is important to recognize, as it is frequently associated with comorbid anxiety and depressive disorders, heightened vulnerability to obsessive-compulsive symptomatology, and reduced |

| | | |
|---|---|---|
| disorder | under continuant unit → sdc unit → realizable unit → disposition unit → PersonCenteredDisposition → PsychologicalDisposition, as the disorder embodies a long-term dispositional tendency that manifests in rigid perfectionism, excessive conscientiousness, and inflexibility in social and personal contexts. This ontological positioning differentiates it from processes, which would capture the contingent unfolding of particular symptomatic episodes, and from states, which would describe temporary observable conditions. By contrast, anankastic personality disorder is defined by an enduring psychological structure that underlies and shapes the probability of certain forms of thought and behavior across contexts and over time. Classifying it as a PsychologicalDisposition therefore ensures ontological precision by recognizing its nature as a realizable but enduring mental predisposition rather than a transient occurrence. | treatment flexibility due to entrenched cognitive and behavioral patterns. The presence and severity of this diagnosis provide interpretive value by indicating a long-standing predisposition toward maladaptive control strategies, impaired adaptability, and elevated risk of stress-induced decompensation, while its absence or effective management may signify improved cognitive flexibility, relational functioning, and treatment responsiveness. Thus, this datapoint functions as a key historical marker linking enduring personality structure to current psychiatric presentation, informing diagnostic formulation, therapeutic planning, and prognosis in psychiatric care. |
| **Previous Diagnoses:**<br><br>Harmful Use of Alcohol | The datapoint is classified in the Meta Mesh Ontology under ClinicalDisposition because it designates an enduring, person-centered tendency that predisposes an individual to recurrent harmful patterns of alcohol consumption and their associated clinical consequences. Within the BFO/MMO hierarchy, this corresponds to continuant unit → sdc unit → realizable unit → disposition unit → PersonCenteredDisposition → ClinicalDisposition, as the condition represents a realizable but stable disposition manifesting in harmful health outcomes when the relevant circumstances - alcohol intake - are present. This classification distinguishes it from processes, which would denote the actual acts of drinking or episodes of intoxication, and from transient states, which would describe short-term physiological or psychological conditions. By recognizing "Harmful Use of Alcohol" as a ClinicalDisposition, the ontology preserves the correct ontological level of abstraction: it captures the enduring clinical vulnerability and health-related predisposition rather than any specific occurrence, thereby aligning with both the medical semantics of ICD-10 and the ontological principles of BFO. | It signifies a maladaptive pattern of alcohol consumption that leads to psychological, social, or physical harm, thereby directly influencing mental health outcomes and treatment trajectories. Clinically, a history of harmful alcohol use is critical to assess, as it is strongly associated with depressive and anxiety disorders, personality pathology, and increased risk of suicidal behavior, while also complicating the course and management of comorbid psychiatric conditions. The presence and severity of this diagnosis provide interpretive value by indicating maladaptive coping mechanisms, heightened vulnerability to relapse, and potential barriers to treatment adherence, whereas its absence or remission may reflect improved self-regulation, enhanced resilience, and greater likelihood of positive therapeutic outcomes. Thus, this datapoint functions as a vitalmarker linking substance-related harm to psychiatric morbidity, informing diagnostic formulation, prognosis, and the integration of addiction-focused strategies into comprehensive psychiatric care. |
| **Previous** | The datapoint is classified in the Meta Mesh Ontology under PsychologicalDisposition because | It reflects a chronic pattern of multiple, recurrent, and medically unexplained physical symptoms that lead to |

| | | |
|---|---|---|
| **Diagnoses:** Somatization Disorder | it represents an enduring, person-centered tendency to manifest psychological distress through recurrent somatic symptoms that cannot be fully explained by identifiable medical conditions. Within the BFO/MMO hierarchy, this corresponds to continuant unit → sdc unit → realizable unit → disposition unit → PersonCenteredDisposition → PsychologicalDisposition, as the disorder reflects a stable disposition inherent in the individual that predisposes them to particular symptom patterns over time. This classification differentiates it from processes, which would describe the episodic occurrences of somatic complaints or clinical encounters, and from transient states, which would only capture momentary manifestations of distress. By situating "Somatization Disorder" as a PsychologicalDisposition, the ontology preserves the appropriate level of abstraction by identifying the enduring psychological predisposition that underlies recurrent somatic presentations, thereby aligning the diagnostic semantics of ICD-10 with the ontological principles of BFO. | significant distress and functional impairment, representing a complex interface between psychological processes and bodily experience. Clinically, a history of somatization disorder is important to consider, as it is strongly associated with depressive and anxiety disorders, trauma-related conditions, and personality pathology, and it frequently complicates psychiatric treatment through high healthcare utilization, reduced trust in medical providers, and resistance to psychological explanations. The presence and severity of this diagnosis provide interpretive value by indicating a persistent vulnerability to somatic expression of psychological distress, potential challenges in therapeutic engagement, and heightened risk of comorbid mood and anxiety pathology, while its absence or successful management may suggest greater psychological insight, adaptive coping, and improved treatment responsiveness. Thus, this datapoint functions as a key historical marker linking somatic symptomatology to psychiatric morbidity, guiding diagnostic formulation and informing integrative treatment approaches that address both mind and body. |

**Table 5.** *An overview of all data points included in the use case is provided, along with two key dimensions of justification: (1) the ontological rationale for the classification of each data point within the Meta Mesh Ontology (MMO) and its alignment with the appropriate Core Domain Framework (CDF) class, and (2) the clinical-psychiatric relevance of the data point. The first column lists the specific data point, the second explains its semantic assignment in the MMO/CDF structure, and the third outlines its relevance within psychiatric assessment or care. This structured mapping supports transparent integration of heterogeneous knowledge elements into a unified ontology-based mental health model.*
